# Antibacterial Treatment and Outcomes in Adults With Virus-Positive Community-Acquired Pneumonia

**DOI:** 10.64898/2026.08.19.26360846

**Authors:** Mayar Al Mohajer, Kasim Allel, David Slusky, David Nix, Catia Nicodemo

## Abstract

**Rationale:** Guidelines disagree on antibacterial treatment for adults with community-acquired pneumonia and a positive respiratory viral test, particularly hospitalized patients and outpatients with comorbidities.

**Objectives:** To estimate associations between antibacterial treatment selected for community-acquired pneumonia and outcomes in adults with virus-positive, imaging-evaluated nonsevere pneumonia.

**Methods:** We conducted a retrospective multicenter study using Epic Cosmos data from 2016–2025. Hospitalized patients treated empirically by 24 hours were compared by continuation during hours 24–48; outpatients were compared by prescription at emergency-department discharge. Analyses were stratified by guideline-defined comorbidity and used propensity-score overlap weighting with source-cluster bootstrap confidence intervals. Exploratory analyses assessed respiratory virus, antiviral treatment, antibacterial class, and outpatient timing.

**Measurements and Main Results:** The cohort included 376,320 adults: 275,604 inpatients and 100,716 outpatients. Inpatients who continued treatment had higher 30-day adverse-event risk without guideline comorbidity (adjusted risk difference, 1.70 percentage points; 95% confidence interval, 0.80–2.39) and with guideline comorbidity (2.56; 1.88–3.14), and longer post-landmark stay (adjusted mean ratios, 1.14 and 1.08). Exploratory class-specific analyses showed the largest adverse-event and mortality associations with broad therapy targeting resistant staphylococci or *Pseudomonas*; macrolide-containing and other atypical coverage showed no consistent adverse signal. Outpatient prescribing was associated with lower risks, but care-transition and residual confounding remained.

**Conclusions:** Continued inpatient therapy after the empiric period showed no evidence of benefit and was associated with worse observed outcomes. Outpatient associations favored prescribing but remained vulnerable to care-transition and residual confounding.

## Introduction

Community-acquired pneumonia (CAP) remains a frequent reason for adult emergency care and hospitalization, and respiratory viruses account for a substantial proportion of microbiologically defined cases (1). Rapid molecular assays identify viral pathogens early, but a positive result does not exclude bacterial coinfection, whose frequency varies by virus and context (1, 2). Empiric antibacterials may be lifesaving when bacterial pneumonia is present, yet unnecessary exposure causes drug-related adverse events, *Clostridioides difficile* infection, and antimicrobial-resistance selection (3).

This uncertainty has produced a guideline disagreement. The updated American Thoracic Society (ATS) guideline suggests withholding empiric antibacterials only for virus-positive outpatients without comorbidities, but suggests CAP-active therapy for outpatients with comorbidities and for patients hospitalized with nonsevere CAP; both recommendations are conditional and based on very low-quality evidence (4). The Infectious Diseases Society of America (IDSA) instead favors individualized withholding or early discontinuation when bacterial pneumonia is unlikely (5). The ATS review identified no direct comparative study after viral identification (4).

Recent observational studies found no clinically meaningful benefit of early CAP-directed therapy in nonsevere COVID-19 and similar outcomes with 0–2 versus 5–7 treatment days among virus-positive hospitalized adults (6, 7). Prior work has not addressed outpatient discharge prescribing, modification by guideline comorbidity or viral pathogen, or whether inpatient associations differ by antibacterial class. We therefore evaluated outpatient prescribing and inpatient continuation after early empiric therapy in a large multicenter cohort of adults with virus-positive, imaging-evaluated CAP, with exploratory virus-specific, antiviral, and class-specific analyses.

## Methods

### Study design and data source

We conducted a retrospective multicenter comparative-effectiveness study using deidentified Epic Cosmos electronic health record data from January 1, 2016, through December 31, 2025. The study addressed virus-positive CAP scenarios at the center of the ATS–IDSA disagreement: outpatient discharge prescribing and inpatient continuation after empiric therapy (4, 5). The 2019 ATS/IDSA framework informed pneumonia and comorbidity definitions (8), and reporting followed the Strengthening the Reporting of Observational Studies in Epidemiology statement (9). Sites contributed fully usable months; only the first eligible episode per patient was retained.

### Ethics and data governance

This analysis used deidentified Epic Cosmos data accessed under institutional permissions and did not constitute human subjects research; informed consent or institutional review board approval was not required under applicable institutional policy. Data were analyzed only as aggregate outputs.

### Cohort construction

We identified adults aged 18 years or older with a complete US emergency-department (ED) or inpatient encounter containing an ICD-10-CM J12–J18 pneumonia diagnosis. Eligibility required a clinically relevant pneumonia diagnosis, a positive respiratory viral molecular or antigen result linked to the encounter, and chest radiography or computed tomography before treatment assignment. Viral results had to be available by ED departure for outpatients and within 48 hours of admission for inpatients. Because report text was not uniformly available, pneumonia was imaging-evaluated but not independently radiologist-adjudicated.

We excluded urgent-care encounters; definite bacterial-pneumonia diagnoses or positive atypical-bacterial results available by assignment; major immunocompromise; high-specificity bacterial laboratory evidence; competing indications for systemic antibacterials; and severe disease before assignment. Severe disease was an intensive care unit stay, airway placement, definite ventilatory support, or intravenous or intraosseous vasopressor use. Observable minor severity features were adjusted for because several ATS/IDSA criteria were not consistently measurable.

### Treatment strategies and clinical strata

For inpatients, the qualifying positive viral result defined time zero. The treatment decision occurred 24 hours later, exposure was assessed during hours 24–48, and follow-up began at the 48-hour landmark. All primary inpatient episodes had received empiric CAP-active therapy by 24 hours and remained alive and hospitalized through the landmark. The comparison was CAP-active antibacterial administration continued during hours 24–48 versus no CAP-active administration during that window. For outpatients, the decision time and analysis start were the actual ED departure. The comparison was a CAP-active discharge prescription versus no CAP-active discharge prescription; administration without a discharge prescription was considered indeterminate and excluded.

CAP-active agents were identified from a reviewed medication map of systemic agents plausibly used for CAP; local routes, antiseptics, and nonabsorbed gastrointestinal agents were excluded. Source-year-setting cells with essentially absent setting-relevant medication capture were excluded. Exploratory overlapping class indicators identified atypical coverage, macrolide-containing therapy, doxycycline/tetracycline, respiratory fluoroquinolone, beta-lactam therapy without atypical coverage, and broad therapy active against methicillin-resistant *Staphylococcus aureus* (MRSA) or *Pseudomonas* species.

### Outcomes

Follow-up began at ED departure for outpatients and at the prespecified landmark for inpatients. The outpatient adverse-event composite included all-cause hospitalization within 14 days, ED revisit within 30 days, or death within 30 days. The inpatient composite included unplanned readmission, ED revisit, death, or clinical deterioration within 30 days. Clinical deterioration was new intensive care, advanced airway placement, invasive or noninvasive ventilation, or vasopressor use after the landmark. Components, positive diagnostic *Clostridioides difficile* results within 30 and 90 days, and inpatient post-landmark length of stay were also evaluated.

### Covariates

Propensity models included variables available by treatment assignment: age; sex; race; ethnicity; payer; state; calendar year; respiratory virus; anonymized source; chronic cardiopulmonary, renal, hepatic, neurologic, metabolic, malignant, and behavioral comorbidities; smoking and obesity indicators; baseline respiratory rate, oxygen saturation, systolic blood pressure, temperature, white blood cell count, platelet count, blood urea nitrogen; and an indicator for three or more observable minor severity criteria. Continuous variables were median-imputed, standardized, and modeled with linear and supported quadratic terms plus missingness indicators. Categorical missingness was explicit, rare categories were collapsed, and missing coded binary indicators were treated as absence.

### Statistical analysis

Within each primary stratum, logistic regression estimated treatment propensity from prespecified covariates. Scores were bounded at 0.001–0.999. Overlap weights were 1 minus the propensity score for treated episodes and the propensity score for untreated episodes, targeting patients with the greatest clinical equipoise (10–13). Balance was assessed with absolute standardized mean differences; values below 0.10 were acceptable.

For binary outcomes, we estimated overlap-weighted risks, adjusted risk differences in percentage points, and risk ratios; for length of stay, weighted means, mean differences, and mean ratios. We used 500 source-cluster bootstrap replications with the fitted propensity model held fixed. Two-sided empirical bootstrap P values quantified risk-difference and interaction contrasts. Negative risk differences indicate lower risk with treatment. Analyses were conducted in R version 4.5.1 (R Foundation for Statistical Computing, Vienna, Austria) using the data.table and splines packages.

### Artificial intelligence tools

OpenAI ChatGPT was used for language editing and refinement. All scientific content and final wording were reviewed and approved by the authors.

### Sensitivity, interaction, and exploratory analyses

The inpatient sensitivity shifted assignment to 48 hours, assessed continuation during hours 48–72, and began follow-up at 72 hours. The outpatient care-transition sensitivity excluded hospitalizations beginning within 24 hours from the hospitalization outcome and composite. Treatment-by-comorbidity interaction was the difference between stratum-specific risk differences using paired source-cluster bootstrap intervals. Exploratory inpatient analyses compared continuation versus noncontinuation of each antibacterial-class indicator with separately fitted propensity models; a limited 72-hour replication evaluated any atypical, macrolide-containing, and broad therapy. A separate outpatient sensitivity excluded episodes with captured post-result ED antibacterial administration and repeated the early-transition-restricted outcomes. A formal conditional 24-hour landmark was not estimable because patient-level death timestamps were unavailable.

Respiratory viruses were classified as severe acute respiratory syndrome coronavirus 2 (SARS-CoV-2), influenza, respiratory syncytial virus, rhinovirus/enterovirus, human metapneumovirus, endemic coronavirus, multiple respiratory viruses, or other virus. Exploratory within-virus models required at least 300 episodes, 50 episodes per treatment arm, five source clusters, and, for binary outcomes, 10 events per arm; global heterogeneity used Wald tests derived from paired source-cluster bootstrap covariance. Target-concordant antivirals were summarized and added to selected inpatient propensity models. Antibacterial regimens were described among treated episodes.

## Results

### Study cohort and treatment patterns

The primary analysis included 376,320 virus-positive, clinician-diagnosed nonsevere CAP episodes: 275,604 inpatient and 100,716 outpatient episodes from 341 and 272 source clusters, respectively. Guideline comorbidity was present in 82.9% and 50.3%. Among inpatients, 209,682 (76.1%) continued CAP-active therapy during hours 24–48, more often with than without comorbidity (77.1% vs. 70.9%). Among outpatients, 42,850 (42.5%) received a discharge prescription, less often with than without comorbidity (36.7% vs. 48.5%). Treatment declined in 2020–2021 and increased through 2025 (Table 1; Figure 1).

**Figure 1.**
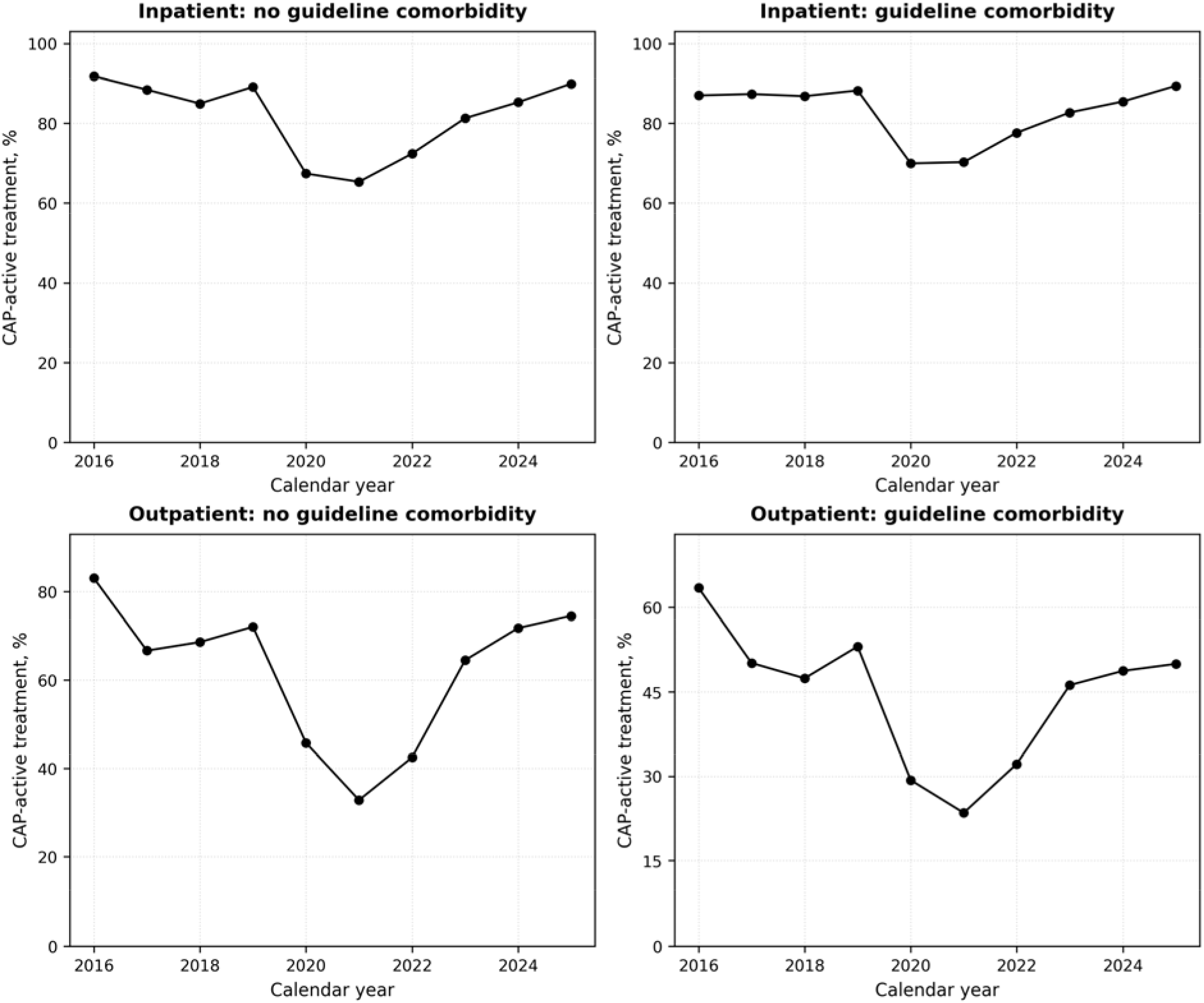
CAP-active antibacterial treatment by calendar year. Annual proportions receiving CAP-active treatment from 2016 through 2025, shown separately for inpatients and outpatients with and without guideline-defined comorbidity. Inpatient treatment denotes continuation during 24–48 hours; outpatient treatment denotes a CAP-active prescription at discharge. CAP = community-acquired pneumonia. Alt text: Four line charts show high antibacterial treatment in 2016–2019, a marked decline in 2020–2021, and partial recovery through 2025. Inpatient treatment remains higher than outpatient treatment throughout; outpatient treatment is lowest among patients with guideline comorbidity during 2020–2021.

**Table 1.**
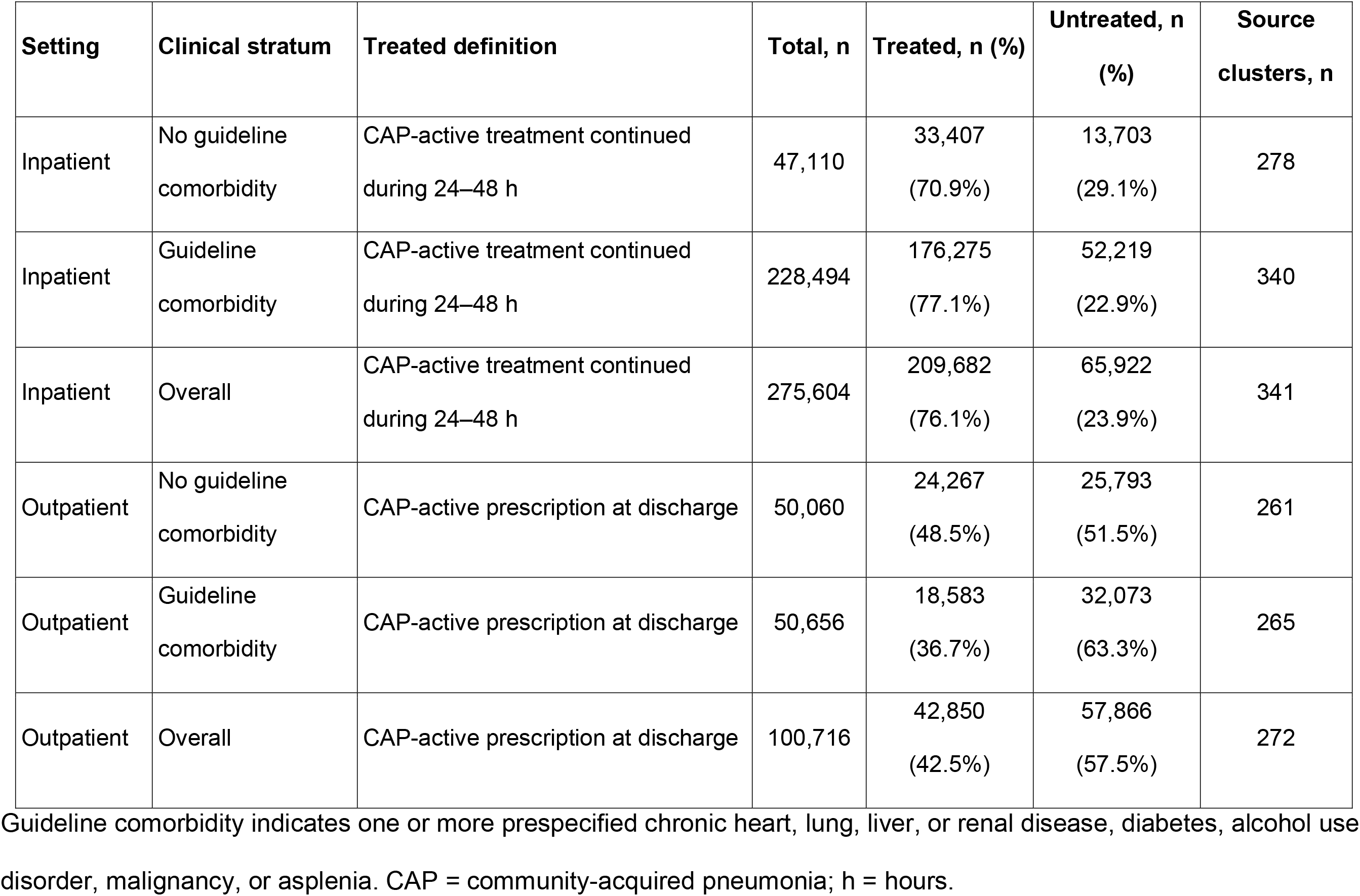
Analysis cohorts and CAP-active antibacterial treatment groups.

| Setting | Clinical stratum | Treated definition | Total, n | Treated, n (%) | Untreated, n (%) | Source clusters, n |
| --- | --- | --- | --- | --- | --- | --- |
| Inpatient | No guideline comorbidity | CAP-active treatment continued during 24–48 h | 47,110 | 33,407 (70.9%) | 13,703 (29.1%) | 278 |
| Inpatient | Guideline comorbidity | CAP-active treatment continued during 24–48 h | 228,494 | 176,275 (77.1%) | 52,219 (22.9%) | 340 |
| Inpatient | Overall | CAP-active treatment continued during 24–48 h | 275,604 | 209,682 (76.1%) | 65,922 (23.9%) | 341 |
| Outpatient | No guideline comorbidity | CAP-active prescription at discharge | 50,060 | 24,267 (48.5%) | 25,793 (51.5%) | 261 |
| Outpatient | Guideline comorbidity | CAP-active prescription at discharge | 50,656 | 18,583 (36.7%) | 32,073 (63.3%) | 265 |
| Outpatient | Overall | CAP-active prescription at discharge | 100,716 | 42,850 (42.5%) | 57,866 (57.5%) | 272 |

Guideline comorbidity indicates one or more prespecified chronic heart, lung, liver, or renal disease, diabetes, alcohol use disorder, malignancy, or asplenia. CAP = community-acquired pneumonia; h = hours.

### Covariate balance

Overlap weighting reduced the maximum absolute standardized mean difference from 0.419–0.600 before weighting to less than 0.001 in every primary stratum; no covariate exceeded 0.10 (Table S1 and Figure S1 in the Online Supplement; Figure 2).

**Figure 2.**
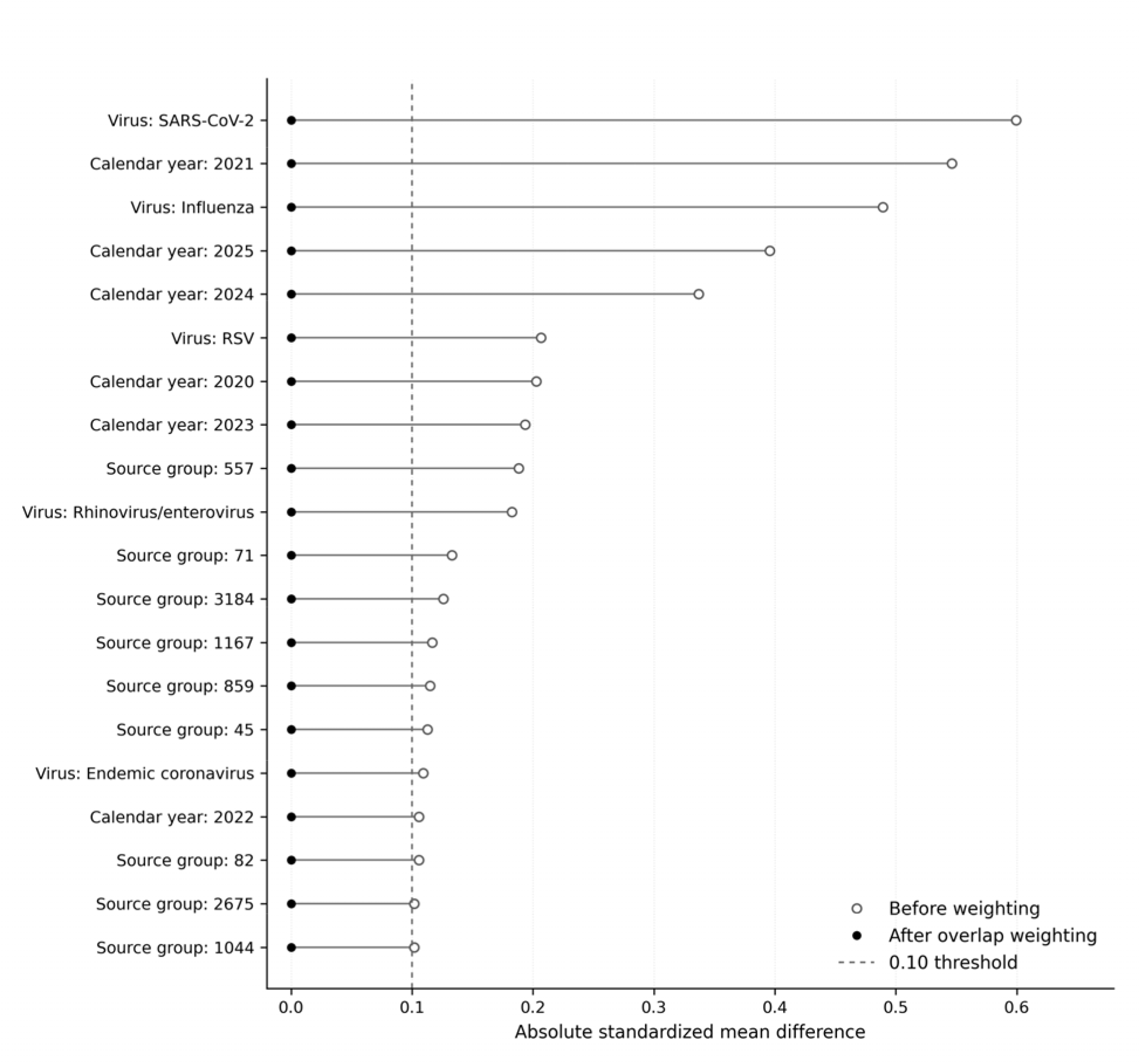
Covariate balance after overlap weighting. The 20 covariate indicators with the largest absolute standardized mean differences before weighting are shown. Open circles indicate values before weighting, filled circles indicate values after overlap weighting, and the dashed line marks the prespecified 0.10 balance threshold. Alt text: A Love plot shows substantial baseline imbalance for viral target, calendar year, and several source groups before weighting. All filled circles after overlap weighting lie essentially at zero and below the 0.10 threshold.

### Inpatient treatment continuation and outcomes

Among inpatients without guideline comorbidity, the adjusted weighted risk of any adverse clinical event within 30 days was 18.24% with continued treatment and 16.54% without continuation (adjusted risk difference, 1.70 percentage points; 95% confidence interval [CI], 0.80 to 2.39; risk ratio, 1.10; 95% CI, 1.05 to 1.15). Among those with guideline comorbidity, the corresponding risks were 33.15% and 30.59% (adjusted risk difference, 2.56 percentage points; 95% CI, 1.88 to 3.14; risk ratio, 1.08; 95% CI, 1.06 to 1.10) (Table 2; Figure 3, panel A).

**Figure 3.**
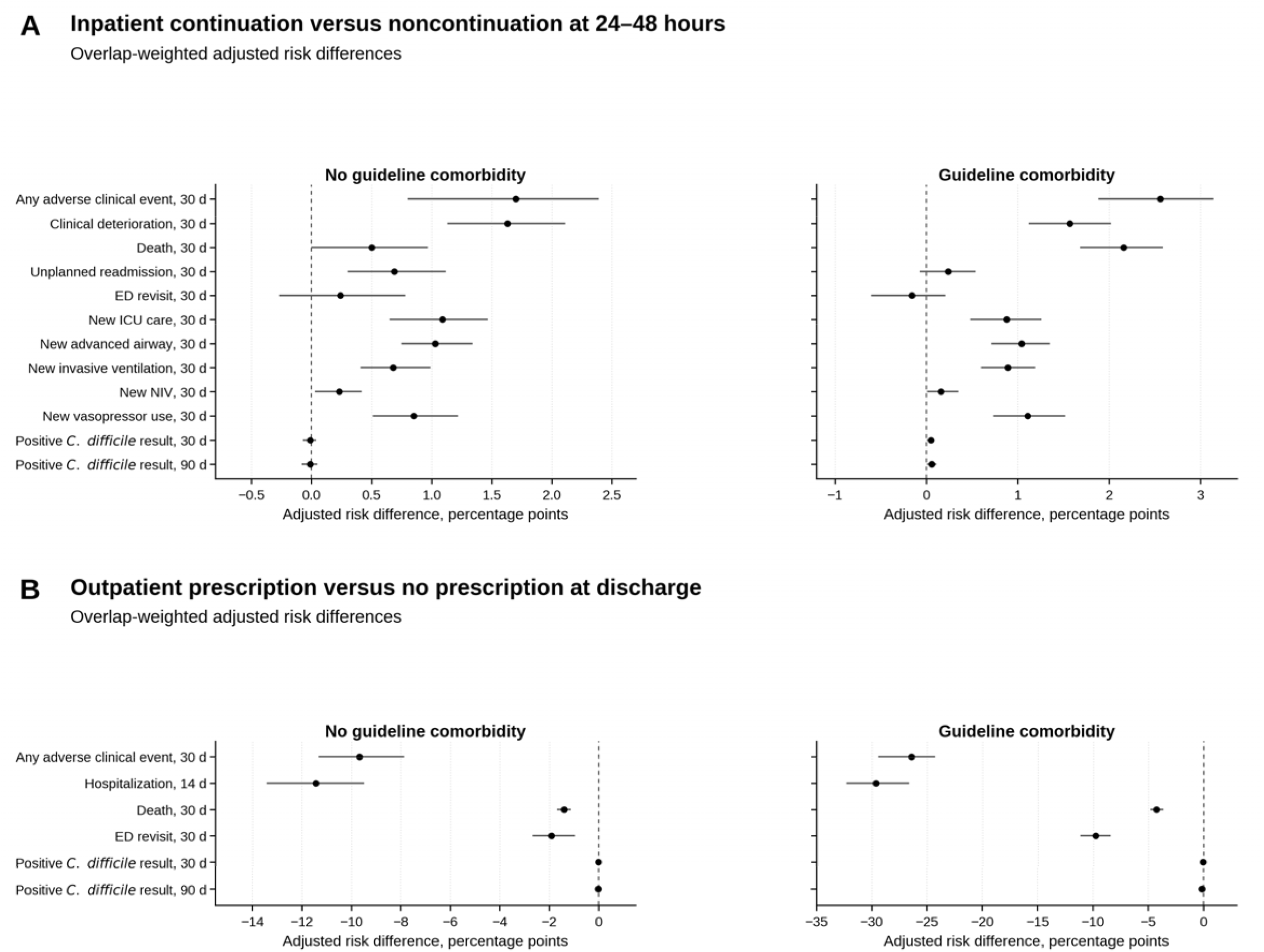
Primary overlap-weighted binary outcome estimates. Panel A compares inpatient continuation versus noncontinuation of CAP-active treatment during 24–48 hours. Panel B compares outpatient CAP-active prescription versus no prescription at discharge. Points are adjusted risk differences and horizontal lines are 95% source-cluster bootstrap confidence intervals. Positive values indicate higher risk with treatment; negative values indicate lower risk with treatment. ED = emergency department; ICU = intensive care unit; NIV = noninvasive ventilation. Alt text: Panel A shows mostly positive adjusted risk differences with inpatient continuation, including adverse events, deterioration, death, and escalation of care. Panel B shows negative risk differences with outpatient prescription, most prominently for hospitalization and the composite adverse-event outcome.

**Table 2.**
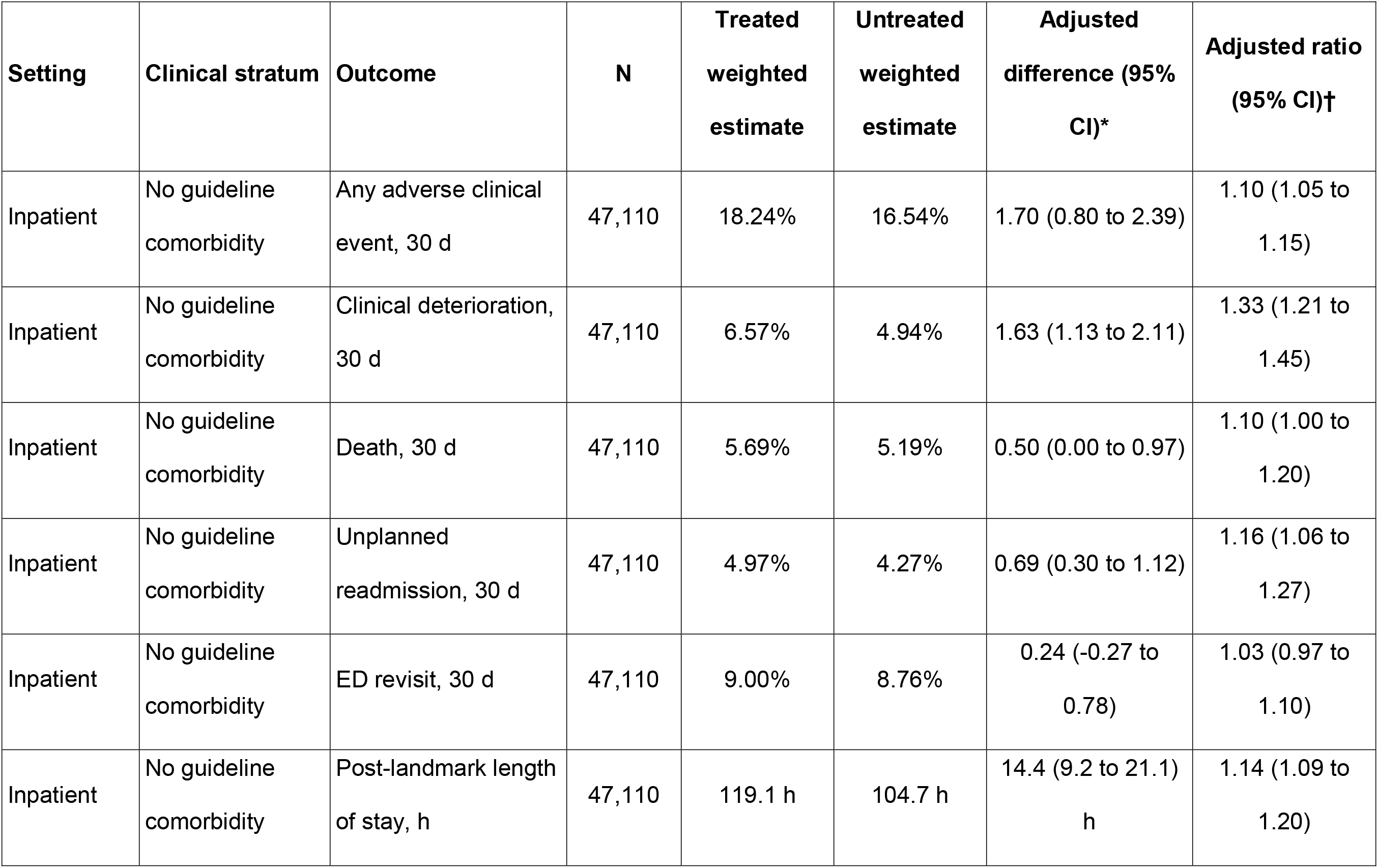
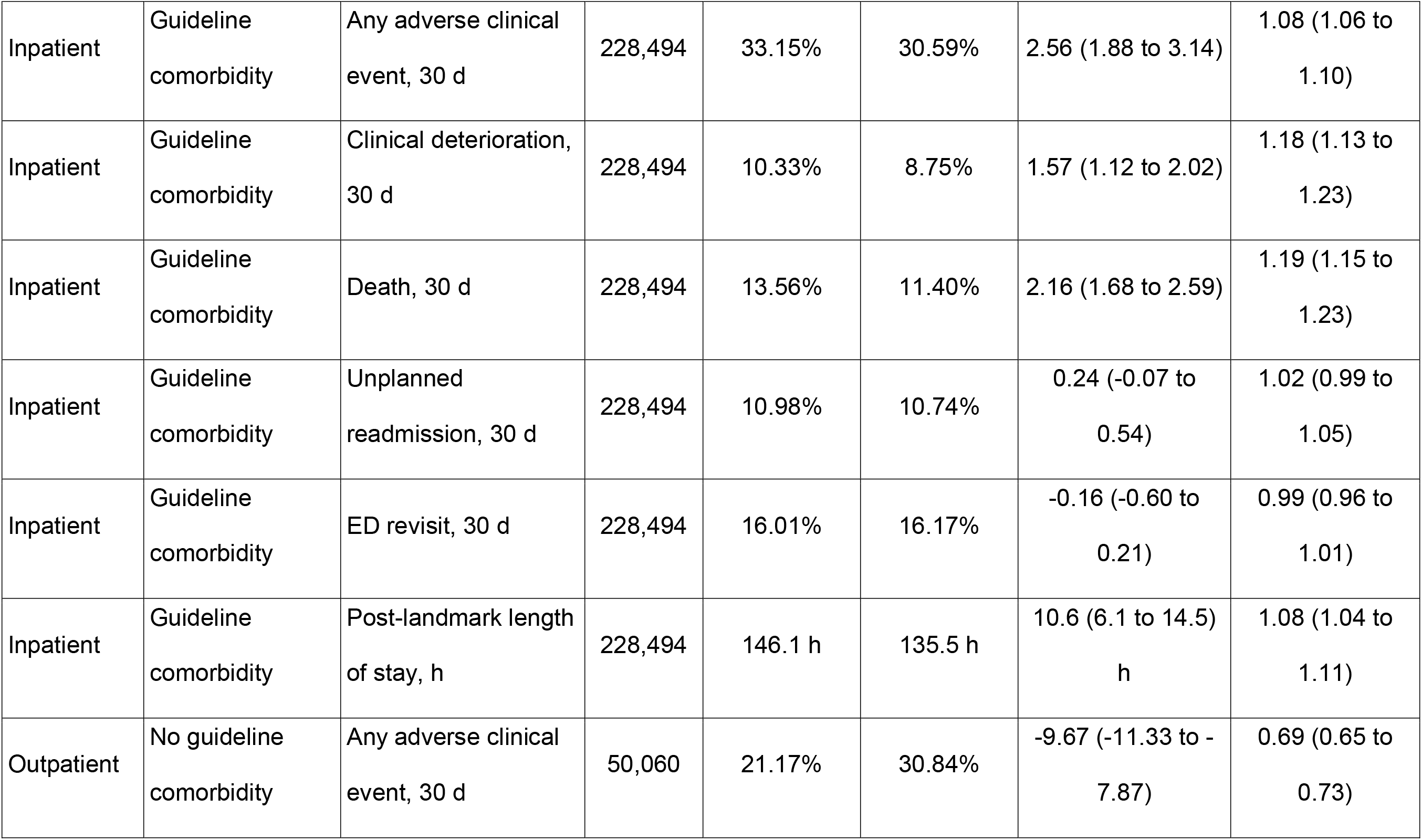
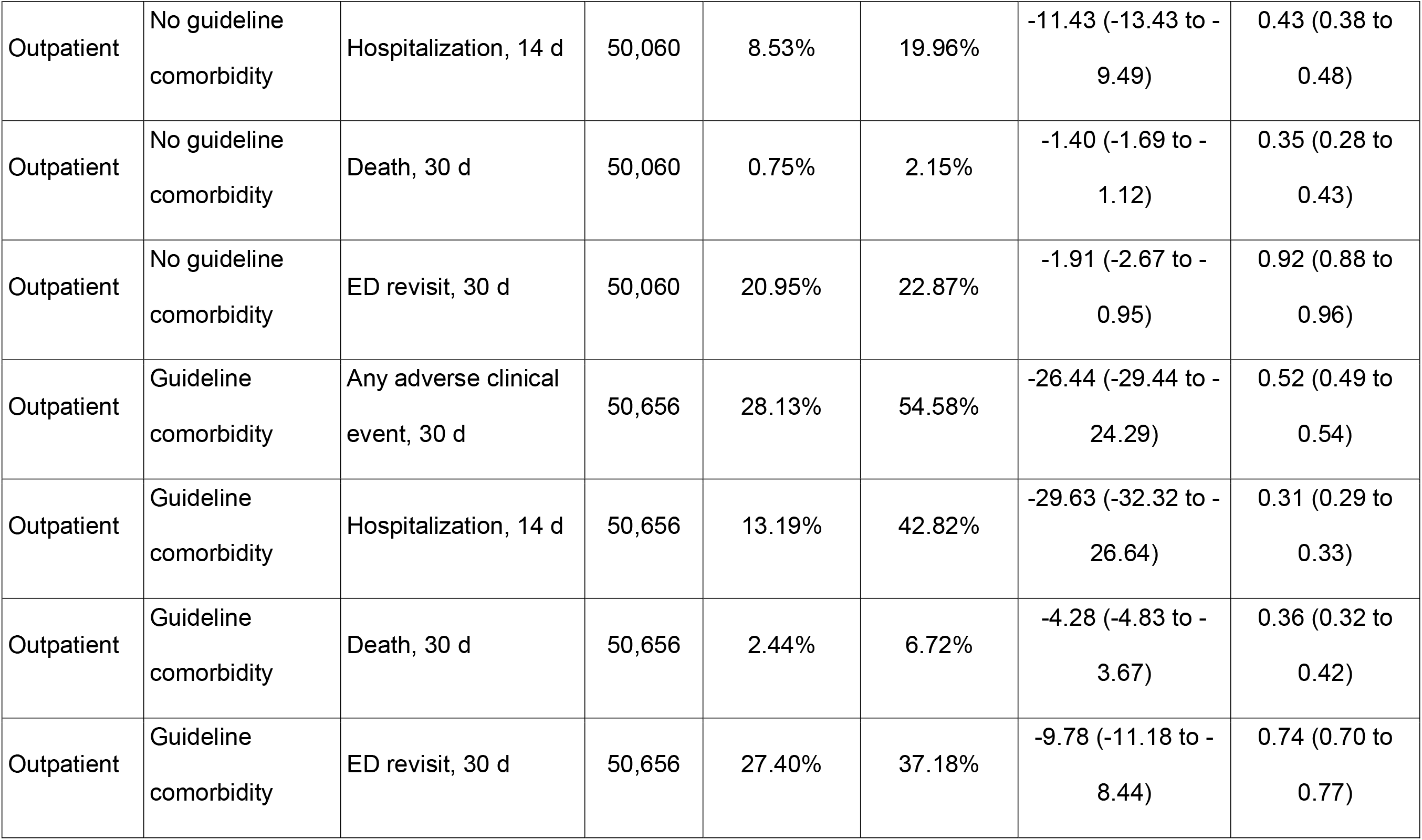
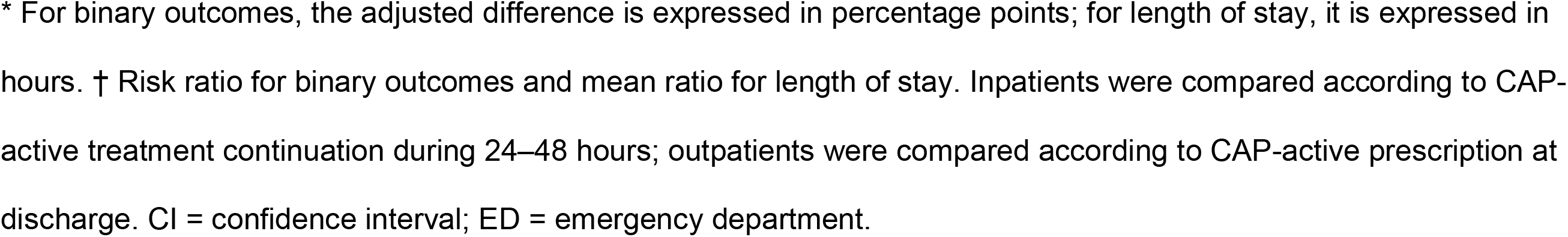
Primary overlap-weighted outcome estimates by setting and guideline-comorbidity stratum.

Continuation was associated with higher clinical-deterioration risk by 1.63 percentage points (95% CI, 1.13 to 2.11) without and 1.57 (1.12 to 2.02) with comorbidity. Death differences were 0.50 (0.00 to 0.97) and 2.16 (1.68 to 2.59), respectively. New intensive care, airway or ventilatory support, and vasopressor use were also more frequent (Table S2 in the Online Supplement). Post-landmark stay was longer by 14.4 hours (9.2 to 21.1) and 10.6 hours (6.1 to 14.5); mean ratios were 1.14 and 1.08 (Figure 4).

**Figure 4.**
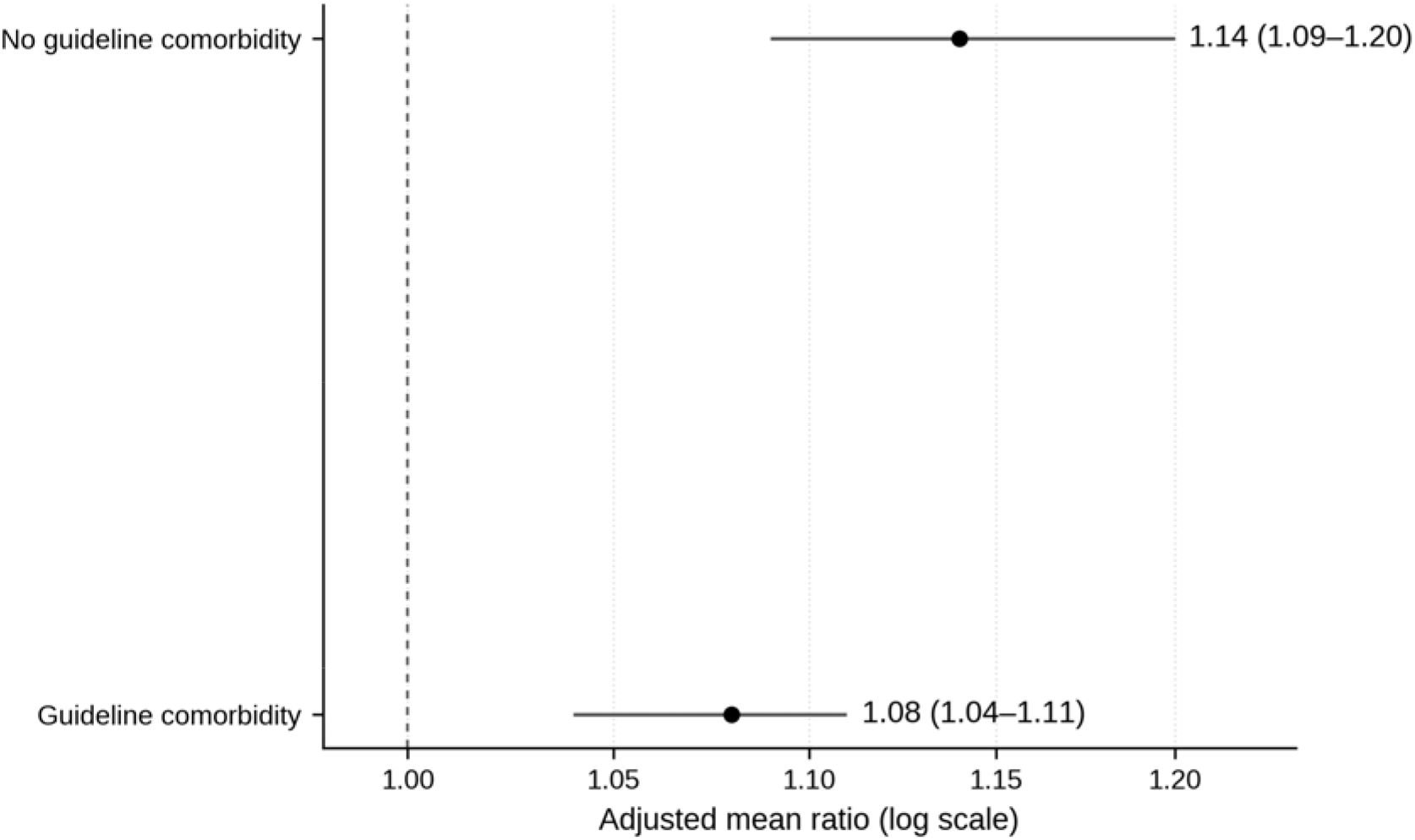
Post-landmark length of stay after overlap weighting. Adjusted mean ratios compare inpatients with CAP-active treatment continued during 24–48 hours with those without continuation. Ratios greater than 1 indicate longer post-landmark length of stay with continuation. Alt text: Two point estimates exceed 1. Continued treatment is associated with a mean ratio of 1.14 in patients without guideline comorbidity and 1.08 in patients with guideline comorbidity; both confidence intervals exclude 1.

### Outpatient prescription and outcomes

In the primary outpatient analysis, discharge prescription was associated with lower risk in both strata. Without comorbidity, adjusted risk differences were -9.67 percentage points (95% CI, -11.33 to -7.87) for the composite and -11.43 (-13.43 to -9.49) for hospitalization. With comorbidity, they were -26.44 (-29.44 to -24.29) and -29.63 (-32.32 to -26.64), respectively. Death and ED revisit were also less frequent (Table 2; Figure 3, panel B).

Immediate hospital transitions were more frequent without a prescription: hospitalization within 0–6 hours occurred in 9.05% versus 0.10% without comorbidity and 24.36% versus 0.38% with comorbidity (Table S3 in the Online Supplement).

Excluding hospitalizations within 24 hours attenuated adjusted differences for the composite to -3.17 and -13.38 percentage points and for hospitalization to -2.05 and - 5.87, respectively (Table 3).

**Table 3.**
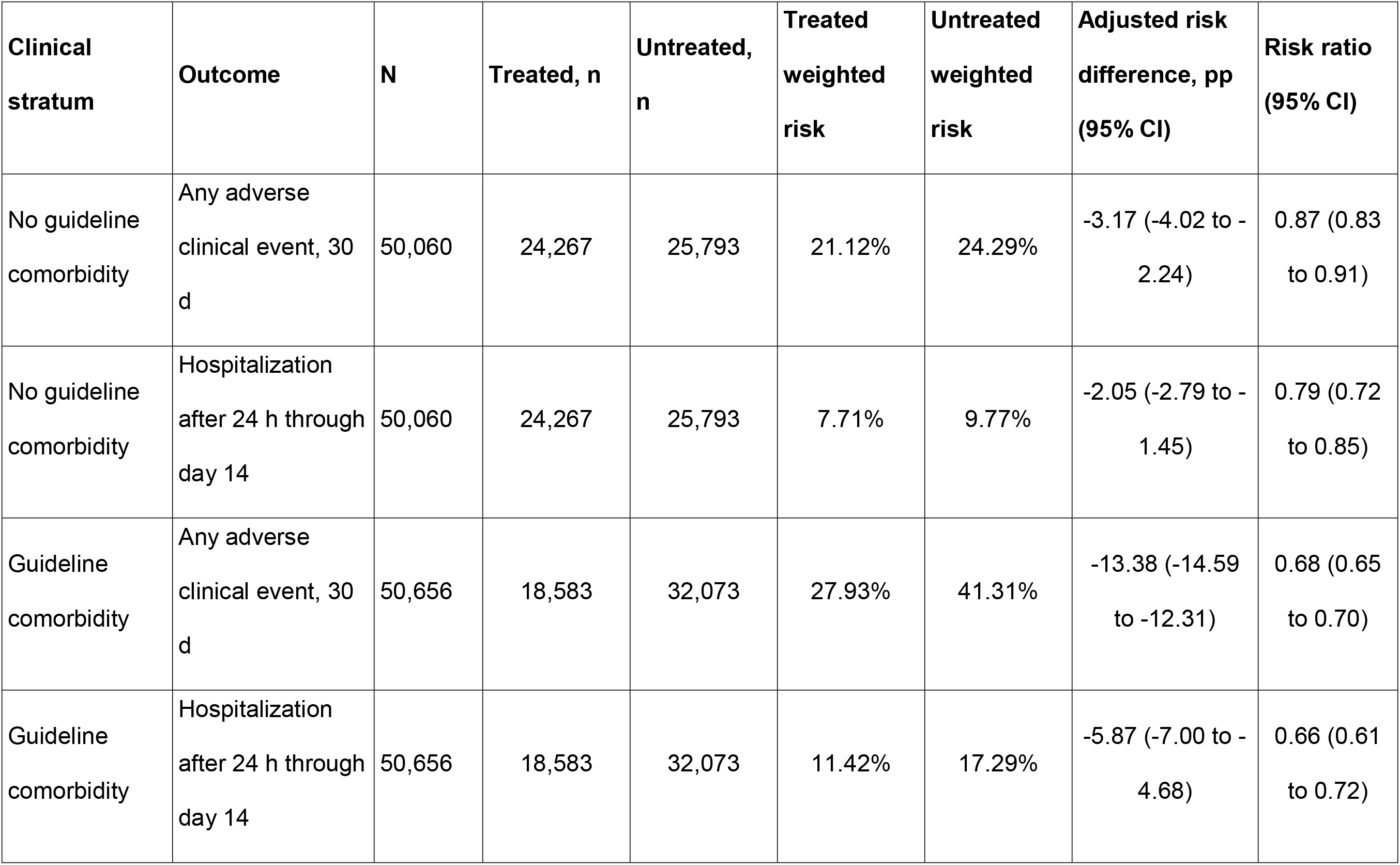
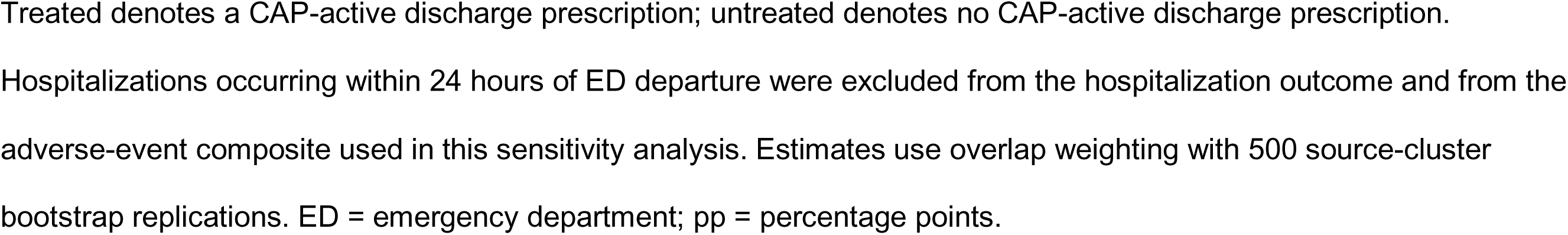
Outpatient 24-hour care-transition sensitivity analysis.

### Sensitivity, interaction, and exploratory analyses

At 72 hours, continuation remained associated with higher composite risk by 1.79 percentage points (95% CI, 0.90 to 2.53) without and 2.30 (1.69 to 3.02) with comorbidity; post-landmark stay remained longer by 10.4 and 11.0 hours (Table S4 in the Online Supplement).

The inpatient treatment-by-comorbidity interaction was 0.87 percentage points (95% CI, -0.04 to 1.80; P = .064) for the composite and 1.66 (0.94 to 2.35; P < .004) for death. Outpatient 24-hour sensitivity interactions were -10.21 (P < .004) for the composite and -3.82 (P < .004) for hospitalization (Table 4).

**Table 4.** Treatment-by-guideline-comorbidity interaction analyses.

| **Analysis** | **Outcome** | **Risk difference in patients without guideline comorbidity, pp (95% CI)** | **Risk difference in patients with guideline comorbidity, pp (95% CI)** | **Interaction risk difference, pp (95% CI)** | **Bootstrap *P* value** |
| --- | --- | --- | --- | --- | --- |
| Primary inpatient | Any adverse clinical event, 30 d | 1.70 (0.81 to 2.51) | 2.56 (1.96 to 3.26) | 0.87 (-0.04 to 1.80) | .064 |
| Primary inpatient | Clinical deterioration, 30 d | 1.63 (1.18 to 2.02) | 1.57 (1.08 to 2.04) | -0.05 (-0.65 to 0.59) | .964 |
| Primary inpatient | Death, 30 d | 0.50 (-0.14 to 0.96) | 2.16 (1.64 to 2.66) | 1.66 (0.94 to 2.35) | <.004 |
| Primary inpatient | ED revisit, 30 d | 0.24 (-0.35 to 0.77) | -0.16 (-0.60 to 0.22) | -0.41 (-1.05 to 0.30) | .216 |
| Primary inpatient | Unplanned readmission, 30 d | 0.69 (0.27 to 1.13) | 0.24 (-0.09 to 0.55) | -0.45 (-0.97 to 0.00) | .056 |
| Outpatient 24-h transition sensitivity | Any adverse clinical event, 30 d | -3.17 (-4.05 to -2.34) | -13.38 (-14.57 to -12.24) | -10.21 (-11.30 to -9.02) | <.004 |
| Outpatient 24-h transition sensitivity | Hospitalization after 24 h through day 14 | -2.05 (-2.69 to -1.48) | -5.87 (-6.96 to -4.68) | -3.82 (-4.82 to -2.76) | <.004 |

The interaction is the risk-difference contrast between the guideline-comorbidity and no-guideline-comorbidity strata. *P*
values are from paired source-cluster bootstrap resampling with 500 replications; an empirical value of zero is reported as
*P* < .004. Stratum-specific confidence intervals in this table were generated from the paired bootstrap replicates used for
the interaction contrast and may therefore differ from the marginal intervals in Tables 2 and 3. CI = confidence interval; ED
= emergency department; pp = percentage points.

### Viral-pathogen, antiviral, and regimen analyses

Severe acute respiratory syndrome coronavirus 2 predominated, accounting for 85.4% of inpatients without guideline comorbidity, 70.6% of inpatients with comorbidity, 74.0% of outpatients without comorbidity, and 72.3% of outpatients with comorbidity.

Influenza accounted for 7.1%–15.6%; other single viruses were less common (Table S5 and Figure S2 in the Online Supplement).

Primary inpatient adverse-event associations varied by virus (global P = .0016 without comorbidity and P < .004 with comorbidity). Risk differences were positive for SARS-CoV-2 (2.02 and 3.57 percentage points) but neutral or inverse for influenza and most other viruses; mortality heterogeneity with comorbidity was confined to SARS-CoV-2. The 72-hour analysis showed the same directional pattern. Outpatient 24-hour sensitivity estimates were generally negative across viruses. Target-concordant antiviral adjustment did not materially alter selected inpatient estimates, although antiviral and antibacterial prescriptions were concurrent for outpatients; regimen distributions varied by setting and virus (Tables S6–S10 and Figures S3–S4 in the Online Supplement).

### Antibacterial-class and outpatient bias analyses

Class-specific inpatient analyses did not identify atypical or macrolide-containing therapy as the source of the adverse associations. Broad anti-MRSA/antipseudomonal continuation showed the largest primary adverse-event differences: 6.89 percentage points (95% CI, 5.28 to 8.63) without and 7.79 (6.84 to 8.65) with comorbidity; death differences were 3.08 (1.80 to 4.33) and 6.05 (5.23 to 6.81). Beta-lactam continuation without atypical coverage was also associated with higher adverse-event and death risks. Broad-therapy associations persisted at 72 hours, whereas any atypical and macrolide-containing estimates were null or modestly inverse (Table S11 and Figures S5A–S5D in the Online Supplement).

Timing-specific outpatient estimates were most extreme for hospitalizations beginning within 6 hours and approached the null after 72 hours (Figure S6 in the Online Supplement). In the analysis restricted to episodes without captured post-result ED antibacterial administration, associations also persisted after excluding hospitalization within 24 hours. Composite risk differences were -2.24 percentage points (95% CI, -3.20 to -1.44) without and -11.61 (-12.70 to -10.58) with comorbidity; hospitalization after 24 hours differed by -1.53 (-2.14 to -0.90) and -6.02 (-7.10 to -5.02), respectively (Table S12 and Figure S7 in the Online Supplement).

## Discussion

In this multicenter comparative-effectiveness study of 376,320 adults with virus-positive, clinician-diagnosed nonsevere CAP, treatment associations differed sharply by setting. Inpatient continuation through 48 hours was associated with higher adverse-event, clinical-deterioration, escalation-of-care, and longer-stay risks; the mortality association was concentrated in the guideline-comorbidity stratum. Exploratory analyses showed that SARS-CoV-2 contributed strongly to pooled binary-outcome associations and that the largest class-specific associations occurred with beta-lactam therapy without atypical coverage and especially broad anti-MRSA/antipseudomonal therapy, not macrolide-containing or other atypical coverage. Findings persisted at 72 hours. Outpatient prescribing was associated with lower risks, including after excluding captured post-result ED administration, but care-transition and residual confounding remained.

The inpatient findings align with contemporary studies reporting no meaningful benefit of early CAP-directed therapy in nonsevere COVID-19 or of 5–7 versus 0–2 treatment days in virus-positive possible CAP (6, 7). Our study extends those observations across multiple viruses, a larger source population, decision-time exclusions, and landmark analyses. Residual confounding remains likely because clinicians may continue therapy for subtle instability not captured in structured data; antibacterial harm may also contribute (3). The class-specific results argue against a macrolide-specific mechanism. Broad therapy and beta-lactams without atypical coverage may instead mark persistent bacterial concern or unmeasured severity. The appropriate interpretation is absence of demonstrated benefit, not proof of causal harm.

Virus-specific results caution against treating the pooled inpatient association as universal. SARS-CoV-2 predominated and had positive, precise estimates, whereas influenza and most other viruses had neutral or inverse estimates. Those inverse estimates do not establish antibacterial benefit because untreated groups were smaller, calendar eras differed, and several confidence intervals were wide. Target-concordant antiviral adjustment did not materially alter selected inpatient estimates, but concurrent antiviral prescribing further limits causal interpretation of outpatient associations.

The findings refine but do not settle the ATS–IDSA disagreement. Our inpatient estimand concerned continuation after initial empiric coverage, not withholding at presentation, and should not be extrapolated to severe CAP, immunocompromise, or bacterial evidence. Within this selected population, the results support structured 24–48-hour reassessment rather than automatic continuation. Guideline comorbidity did not identify a subgroup with apparent benefit for the composite, and the larger mortality association with comorbidity warrants caution because residual severity confounding is plausible. Virus identity may inform reassessment but should not function as an isolated stop-or-treat rule.

The outpatient findings remain less causally secure. Immediate hospitalization was concentrated among nonprescribed patients, indicating that treatment and disposition were closely entangled. Associations attenuated after first-day transitions were removed and persisted after excluding captured post-result ED antibacterial administration, making ED treatment alone an unlikely explanation. However, prescriptions do not establish filling or adherence; concurrent antiviral prescribing, unmeasured clinician judgment, follow-up intensity, and evolving illness may remain. A formal 24-hour conditional landmark could not be constructed. These data neither prove benefit nor demonstrate safe withholding; they identify outpatient treatment, especially with comorbidity, as the key randomized evidence gap.

Diagnostic-stewardship trials support explicit reassessment pathways. Routine viral testing increased brief courses without worse outcomes in ResPOC; viral testing plus procalcitonin reduced discharge prescribing when the algorithm was followed; and a probability-guided trial reduced inpatient days of therapy without a safety signal (14–16). Viral results should therefore be interpreted with clinical trajectory, imaging, biomarkers, microbiology, and competing diagnoses rather than as an isolated stop-or-treat signal.

Strengths include the large 10-year multicenter cohort; separate outpatient and inpatient estimands; decision-aligned landmarks; excellent measured balance; source-cluster resampling; later inpatient and early-transition outpatient sensitivity analyses; and exploratory analyses of viral heterogeneity, antiviral treatment, antibacterial class, and captured ED administration.

Treatment was not randomized, so residual confounding by indication, response, and clinician assessment remains. Pneumonia was code-defined and imaging-evaluated without report text or independent infiltrate confirmation; severity measurement and bacterial testing were incomplete. Virus-specific analyses were exploratory, spanned different calendar eras and overlap populations, and included some small untreated groups. Class-specific indicators were overlapping, and broad therapy likely captured unmeasured bacterial concern. Antiviral exposure reflected documented treatment rather than adherence. Outpatient prescriptions did not establish dispensing or adherence, captured ED administration may be incomplete, and events outside Cosmos organizations may have been missed. Source anonymity limited hospital-level analysis, COVID-era changes may persist despite adjustment, and low *C. difficile* event rates limited safety inference.

Clinically, these findings support cautious, virus-informed reassessment. For stable inpatients with nonsevere CAP, a positive viral assay, no competing indication, and no emerging bacterial evidence, continued therapy after 24–48 hours should be reconsidered rather than automatic; persistent bacterial concern or instability may justify treatment. Non-SARS-CoV-2 estimates were not uniformly adverse and should not justify automatic discontinuation. Outpatient decisions should remain individualized with reliable follow-up and return precautions. Pragmatic randomized trials should examine setting, virus, comorbidity, antibacterial class, antiviral use, and treatment receipt.

## Conclusion

Among selected adults with virus-positive nonsevere CAP, continued inpatient CAP-active therapy showed no evidence of benefit and was associated with worse observed outcomes. Exploratory associations were most consistent for SARS-CoV-2 and for broad anti-MRSA/antipseudomonal or beta-lactam therapy without atypical coverage; non-SARS-CoV-2 and class-specific estimates remain vulnerable to residual confounding. Outpatient prescribing remained associated with lower risks after additional sensitivity analyses, but care-transition, treatment-receipt, concurrent-treatment, and residual confounding preclude causal interpretation. These findings support structured inpatient reassessment and randomized outpatient evidence.

## Impact Statement

Among selected adults with virus-positive nonsevere community-acquired pneumonia, continued inpatient antibacterial therapy selected for community-acquired pneumonia was not associated with improved outcomes; the largest exploratory adverse associations occurred with broad-spectrum therapy targeting resistant staphylococci or *Pseudomonas* and with beta-lactam therapy without atypical coverage. Outpatient associations persisted after excluding captured emergency-department administration but remained vulnerable to care-transition and residual confounding, supporting structured inpatient reassessment and randomized outpatient evidence.

## Author Contributions

M.A.M.: Conceptualization, Methodology, Data curation, Formal analysis, Investigation, Writing - original draft, Writing - review and editing. K.A.: Methodology, Validation, Supervision, Writing - review and editing. D.S.: Methodology, Validation, Supervision, Writing - review and editing. D.N.: Clinical interpretation, Validation, Writing - review and editing. C.N.: Methodology, Validation, Supervision, Writing - review and editing.

## Data Availability

The data underlying this article were accessed through Epic Cosmos under institutional permissions and cannot be shared publicly because of data use agreements.

Aggregated outputs supporting the findings are included in the tables and online supplement.

## Artificial Intelligence Disclosure

OpenAI ChatGPT was used for language editing and refinement of the manuscript. All scientific decisions, analyses, interpretations, and final wording were reviewed and approved by the authors, who take full responsibility for the work.

## References

1. Jain S, Self WH, Wunderink RG, Fakhran S, Balk R, Bramley AM, et al. Community-acquired pneumonia requiring hospitalization among U.S. adults. N Engl J Med. 2015;373(5):415–427. doi:10.1056/NEJMoa1500245.

2. Hedberg P, Johansson N, Ternhag A, Abdel-Halim L, Hedlund J, Nauclér P. Bacterial co-infections in community-acquired pneumonia caused by SARS-CoV-2, influenza virus and respiratory syncytial virus. BMC Infect Dis. 2022;22:108. doi:10.1186/s12879-022-07089-9.

3. Tamma PD, Avdic E, Li DX, Dzintars K, Cosgrove SE. Association of adverse events with antibiotic use in hospitalized patients. JAMA Intern Med. 2017;177(9):1308–1315. doi:10.1001/jamainternmed.2017.1938.

4. Jones BE, Ramirez JA, Oren E, Soni NJ, Sullivan LR, Restrepo MI, et al. Diagnosis and management of community-acquired pneumonia: an official American Thoracic Society clinical practice guideline. Am J Respir Crit Care Med. 2026;212(1):24–44. doi:10.1164/rccm.202507-1692ST.

5. Klompas M, Al-Hasan M, Al Mohajer M, Colgrove R, Doron S, File T, et al. Infectious Diseases Society of America (IDSA) position statement: why IDSA did not endorse the community-acquired pneumonia guidelines 2025 update. Clin Infect Dis. 2026;82(4):622–624. doi:10.1093/cid/ciaf625.

6. Pulia MS, Griffin M, Schwei R, Pop-Vicas A, Schulz LT, Shieh MS, et al. Antibiotic treatment in patients hospitalized for nonsevere COVID-19. JAMA Netw Open. 2025;8(5):e2511499. doi:10.1001/jamanetworkopen.2025.11499.

7. Biebelberg B, Chen T, McKenna C, Kanjilal S, Shappell C, Rhee C, et al. Associations between antibiotic use and outcomes in patients hospitalized with community-acquired pneumonia and positive respiratory viral assays. Clin Infect Dis. 2026;82(4):630–638. doi:10.1093/cid/ciaf687.

8. Metlay JP, Waterer GW, Long AC, Anzueto A, Brozek J, Crothers K, et al. Diagnosis and treatment of adults with community-acquired pneumonia. An official clinical practice guideline of the American Thoracic Society and Infectious Diseases Society of America. Am J Respir Crit Care Med. 2019;200(7):e45–e67. doi:10.1164/rccm.201908-1581ST.

9. von Elm E, Altman DG, Egger M, Pocock SJ, Gøtzsche PC, Vandenbroucke JP; STROBE Initiative. The Strengthening the Reporting of Observational Studies in Epidemiology (STROBE) statement: guidelines for reporting observational studies. Lancet. 2007;370(9596):1453–1457. doi:10.1016/S0140-6736(07)61602-X.

10. Li F, Morgan KL, Zaslavsky AM. Balancing covariates via propensity score weighting. J Am Stat Assoc. 2018;113(521):390–400. doi:10.1080/01621459.2016.1260466.

11. Li F, Thomas LE, Li F. Addressing extreme propensity scores via the overlap weights. Am J Epidemiol. 2019;188(1):250–257. doi:10.1093/aje/kwy201.

12. Austin PC, Stuart EA. Moving towards best practice when using inverse probability of treatment weighting (IPTW) using the propensity score to estimate causal treatment effects in observational studies. Stat Med. 2015;34(28):3661–3679. doi:10.1002/sim.6607.

13. Thomas LE, Li F, Pencina MJ. Overlap weighting: a propensity score method that mimics attributes of a randomized clinical trial. JAMA. 2020;323(23):2417–2418. doi:10.1001/jama.2020.7819.

14. Brendish NJ, Malachira AK, Armstrong L, Houghton R, Aitken S, Nyimbili E, et al. Routine molecular point-of-care testing for respiratory viruses in adults presenting to hospital with acute respiratory illness (ResPOC): a pragmatic, open-label, randomised controlled trial. Lancet Respir Med. 2017;5(5):401–411. doi:10.1016/S2213-2600(17)30120-0.

15. Branche AR, Walsh EE, Vargas R, Hulbert B, Formica MA, Baran A, et al. Serum procalcitonin measurement and viral testing to guide antibiotic use for respiratory infections in hospitalized adults: a randomized controlled trial. J Infect Dis. 2015;212(11):1692–1700. doi:10.1093/infdis/jiv252.

16. Baghdadi JD, Harris AD, Pineles L, Al-Shanqeeti S, Palacio D, Charles DW, et al. Using probability of community-acquired pneumonia to tailor antimicrobials among inpatients: a pragmatic, randomized trial. Clin Infect Dis. 2026;82(6):954–961. doi:10.1093/cid/ciag126.

