## Supplement for "Antibacterial Treatment and Outcomes in Adults With Virus-Positive Community-Acquired Pneumonia"

**Online Supplement**

**Supplementary Table S1. Propensity-score overlap and covariate-balance diagnostics (Part A: propensity-score diagnostics)**

| **Analysis** | **Setting** | **Clinical stratum** | **N** | **Source clusters** | **Propensity-score range** | **Propensity score P1/median/P99** |
| --- | --- | --- | --- | --- | --- | --- |
| Primary | Inpatient | No guideline comorbidity | 47,110 | 278 | 0.326–0.995 | 0.385/0.700/0.963 |
| Primary | Inpatient | Guideline comorbidity | 228,494 | 340 | 0.382–0.999 | 0.468/0.779/0.952 |
| Primary | Outpatient | No guideline comorbidity | 50,060 | 261 | 0.099–0.935 | 0.159/0.432/0.884 |
| Primary | Outpatient | Guideline comorbidity | 50,656 | 265 | 0.073–0.838 | 0.101/0.352/0.706 |
| 72-h sensitivity | Inpatient | No guideline comorbidity | 38,381 | 273 | 0.234–0.994 | 0.346/0.663/0.955 |
| 72-h sensitivity | Inpatient | Guideline comorbidity | 191,782 | 339 | 0.336–0.999 | 0.426/0.743/0.940 |

P1 = first percentile; P99 = 99th percentile.

**Supplementary Table S1. Propensity-score overlap and covariate-balance diagnostics (Part B: overlap-weight and covariate-balance diagnostics)**

| **Analysis** | **Setting** | **Clinical stratum** | **Overlap weight min/median/max** | **Maximum \|SMD\| before** | **Maximum \|SMD\| after** | **Covariates ≥0.10 before/after** |
| --- | --- | --- | --- | --- | --- | --- |
| Primary | Inpatient | No guideline comorbidity | 0.005/0.342/0.984 | 0.419 | 2.20e-08 | 13/0 |
| Primary | Inpatient | Guideline comorbidity | 0.001/0.245/0.987 | 0.420 | 2.31e-07 | 12/0 |
| Primary | Outpatient | No guideline comorbidity | 0.065/0.384/0.935 | 0.600 | 1.28e-12 | 12/0 |
| Primary | Outpatient | Guideline comorbidity | 0.073/0.411/0.927 | 0.423 | 4.34e-09 | 12/0 |
| 72-h sensitivity | Inpatient | No guideline comorbidity | 0.006/0.369/0.978 | 0.366 | 3.27e-08 | 10/0 |
| 72-h sensitivity | Inpatient | Guideline comorbidity | 0.001/0.291/0.982 | 0.405 | 2.56e-07 | 12/0 |

All assessed covariate indicators had an absolute standardized mean difference below 0.10 after overlap weighting. SMD = standardized mean difference.

**Supplementary Table S2. Complete primary overlap-weighted effect estimates**

| **Setting** | **Clinical stratum** | **Outcome** | **N** | **Treated weighted estimate** | **Untreated weighted estimate** | **Adjusted difference (95% CI)*** | **Adjusted ratio (95% CI)†** |
| --- | --- | --- | --- | --- | --- | --- | --- |
| Inpatient | No guideline comorbidity | Any adverse clinical event, 30 d | 47,110 | 18.24% | 16.54% | 1.70 (0.80 to 2.39) | 1.10 (1.05 to 1.15) |
| Inpatient | No guideline comorbidity | Clinical deterioration, 30 d | 47,110 | 6.57% | 4.94% | 1.63 (1.13 to 2.11) | 1.33 (1.21 to 1.45) |
| Inpatient | No guideline comorbidity | Death, 30 d | 47,110 | 5.69% | 5.19% | 0.50 (0.00 to 0.97) | 1.10 (1.00 to 1.20) |
| Inpatient | No guideline comorbidity | Unplanned readmission, 30 d | 47,110 | 4.97% | 4.27% | 0.69 (0.30 to 1.12) | 1.16 (1.06 to 1.27) |
| Inpatient | No guideline comorbidity | ED revisit, 30 d | 47,110 | 9.00% | 8.76% | 0.24 (-0.27 to 0.78) | 1.03 (0.97 to 1.10) |
| Inpatient | No guideline comorbidity | New ICU care, 30 d | 47,110 | 4.92% | 3.83% | 1.09 (0.65 to 1.47) | 1.29 (1.16 to 1.40) |
| Inpatient | No guideline comorbidity | New advanced airway, 30 d | 47,110 | 3.15% | 2.12% | 1.03 (0.75 to 1.34) | 1.48 (1.32 to 1.70) |
| Inpatient | No guideline comorbidity | New invasive ventilation, 30 d | 47,110 | 2.14% | 1.46% | 0.68 (0.41 to 0.99) | 1.46 (1.25 to 1.72) |
| Inpatient | No guideline comorbidity | New noninvasive ventilation, 30 d | 47,110 | 0.94% | 0.71% | 0.23 (0.03 to 0.42) | 1.33 (1.03 to 1.68) |
| Inpatient | No guideline comorbidity | New vasopressor use, 30 d | 47,110 | 3.51% | 2.66% | 0.85 (0.51 to 1.22) | 1.32 (1.18 to 1.50) |
| Inpatient | No guideline comorbidity | Positive *C. difficile* result, 30 d | 47,110 | 0.06% | 0.08% | -0.01 (-0.07 to 0.04) | 0.82 (0.38 to 2.22) |
| Inpatient | No guideline comorbidity | Positive *C. difficile* result, 90 d | 47,110 | 0.10% | 0.11% | -0.01 (-0.08 to 0.05) | 0.89 (0.52 to 1.86) |
| Inpatient | No guideline comorbidity | Post-landmark length of stay, h | 47,110 | 119.1 h | 104.7 h | 14.4 (9.2 to 21.1) h | 1.14 (1.09 to 1.20) |
| Inpatient | Guideline comorbidity | Any adverse clinical event, 30 d | 228,494 | 33.15% | 30.59% | 2.56 (1.88 to 3.14) | 1.08 (1.06 to 1.10) |
| Inpatient | Guideline comorbidity | Clinical deterioration, 30 d | 228,494 | 10.33% | 8.75% | 1.57 (1.12 to 2.02) | 1.18 (1.13 to 1.23) |
| Inpatient | Guideline comorbidity | Death, 30 d | 228,494 | 13.56% | 11.40% | 2.16 (1.68 to 2.59) | 1.19 (1.15 to 1.23) |
| Inpatient | Guideline comorbidity | Unplanned readmission, 30 d | 228,494 | 10.98% | 10.74% | 0.24 (-0.07 to 0.54) | 1.02 (0.99 to 1.05) |
| Inpatient | Guideline comorbidity | ED revisit, 30 d | 228,494 | 16.01% | 16.17% | -0.16 (-0.60 to 0.21) | 0.99 (0.96 to 1.01) |
| Inpatient | Guideline comorbidity | New ICU care, 30 d | 228,494 | 7.59% | 6.71% | 0.88 (0.48 to 1.26) | 1.13 (1.07 to 1.19) |
| Inpatient | Guideline comorbidity | New advanced airway, 30 d | 228,494 | 5.04% | 4.00% | 1.04 (0.71 to 1.35) | 1.26 (1.18 to 1.35) |
| Inpatient | Guideline comorbidity | New invasive ventilation, 30 d | 228,494 | 3.57% | 2.69% | 0.89 (0.60 to 1.19) | 1.33 (1.23 to 1.47) |
| Inpatient | Guideline comorbidity | New noninvasive ventilation, 30 d | 228,494 | 1.45% | 1.29% | 0.16 (0.01 to 0.35) | 1.12 (1.01 to 1.28) |
| Inpatient | Guideline comorbidity | New vasopressor use, 30 d | 228,494 | 6.23% | 5.11% | 1.11 (0.73 to 1.52) | 1.22 (1.14 to 1.31) |
| Inpatient | Guideline comorbidity | Positive *C. difficile* result, 30 d | 228,494 | 0.19% | 0.14% | 0.05 (0.01 to 0.09) | 1.38 (1.08 to 1.88) |
| Inpatient | Guideline comorbidity | Positive *C. difficile* result, 90 d | 228,494 | 0.32% | 0.26% | 0.06 (0.01 to 0.11) | 1.24 (1.05 to 1.49) |
| Inpatient | Guideline comorbidity | Post-landmark length of stay, h | 228,494 | 146.1 h | 135.5 h | 10.6 (6.1 to 14.5) h | 1.08 (1.04 to 1.11) |
| Outpatient | No guideline comorbidity | Any adverse clinical event, 30 d | 50,060 | 21.17% | 30.84% | -9.67 (-11.33 to -7.87) | 0.69 (0.65 to 0.73) |
| Outpatient | No guideline comorbidity | Hospitalization, 14 d | 50,060 | 8.53% | 19.96% | -11.43 (-13.43 to -9.49) | 0.43 (0.38 to 0.48) |
| Outpatient | No guideline comorbidity | Death, 30 d | 50,060 | 0.75% | 2.15% | -1.40 (-1.69 to -1.12) | 0.35 (0.28 to 0.43) |
| Outpatient | No guideline comorbidity | ED revisit, 30 d | 50,060 | 20.95% | 22.87% | -1.91 (-2.67 to -0.95) | 0.92 (0.88 to 0.96) |
| Outpatient | No guideline comorbidity | Positive *C. difficile* result, 30 d | 50,060 | 0.01% | 0.02% | -0.02 (-0.04 to 0.01) | 0.36 (not estimable to 1.73) |
| Outpatient | No guideline comorbidity | Positive *C. difficile* result, 90 d | 50,060 | 0.02% | 0.04% | -0.02 (-0.05 to 0.01) | 0.47 (0.08 to 1.40) |
| Outpatient | Guideline comorbidity | Any adverse clinical event, 30 d | 50,656 | 28.13% | 54.58% | -26.44 (-29.44 to -24.29) | 0.52 (0.49 to 0.54) |
| Outpatient | Guideline comorbidity | Hospitalization, 14 d | 50,656 | 13.19% | 42.82% | -29.63 (-32.32 to -26.64) | 0.31 (0.29 to 0.33) |
| Outpatient | Guideline comorbidity | Death, 30 d | 50,656 | 2.44% | 6.72% | -4.28 (-4.83 to -3.67) | 0.36 (0.32 to 0.42) |
| Outpatient | Guideline comorbidity | ED revisit, 30 d | 50,656 | 27.40% | 37.18% | -9.78 (-11.18 to -8.44) | 0.74 (0.70 to 0.77) |
| Outpatient | Guideline comorbidity | Positive *C. difficile* result, 30 d | 50,656 | 0.07% | 0.12% | -0.06 (-0.11 to -0.01) | 0.53 (0.24 to 0.93) |
| Outpatient | Guideline comorbidity | Positive *C. difficile* result, 90 d | 50,656 | 0.10% | 0.22% | -0.13 (-0.19 to -0.05) | 0.43 (0.21 to 0.73) |

* Percentage points for binary outcomes and hours for post-landmark length of stay. † Risk ratio for binary outcomes and mean ratio for length of stay. CI = confidence interval; ED = emergency department; ICU = intensive care unit; NIV = noninvasive ventilation.

**Supplementary Table S3. Timing of hospitalization after emergency-department departure among outpatients**

| **Clinical stratum** | **Discharge treatment strategy** | **Denominator, n** | **0–6 h** | **>6–24 h** | **>24–72 h** | **>72 h–14 d** | **No hospitalization within 14 d** |
| --- | --- | --- | --- | --- | --- | --- | --- |
| No guideline comorbidity | No CAP-active prescription | 25,793 | 9.05% | 1.57% | 5.61% | 5.06% | 78.71% |
| No guideline comorbidity | CAP-active prescription | 24,267 | 0.10% | 0.58% | 2.37% | 3.75% | 93.20% |
| Guideline comorbidity | No CAP-active prescription | 32,073 | 24.36% | 4.17% | 11.25% | 5.90% | 54.33% |
| Guideline comorbidity | CAP-active prescription | 18,583 | 0.38% | 1.32% | 4.88% | 5.55% | 87.87% |

Percentages are calculated within each clinical-stratum and discharge-treatment group. The marked early transition imbalance motivated the prespecified 24-hour sensitivity analysis. CAP = community-acquired pneumonia.

**Supplementary Table S4. Selected inpatient 72-hour sensitivity estimates**

| **Clinical stratum** | **Outcome** | **N** | **Continued, n** | **Not continued, n** | **Continued weighted estimate** | **Not-continued weighted estimate** | **Adjusted difference (95% CI)** | **Adjusted ratio (95% CI)** |
| --- | --- | --- | --- | --- | --- | --- | --- | --- |
| No guideline comorbidity | Any adverse clinical event, 30 d | 38,381 | 25,582 | 12,799 | 18.03% | 16.24% | 1.79 (0.90 to 2.53) | 1.11 (1.05 to 1.16) |
| No guideline comorbidity | Clinical deterioration, 30 d | 38,381 | 25,582 | 12,799 | 6.05% | 4.54% | 1.52 (1.00 to 2.11) | 1.33 (1.20 to 1.47) |
| No guideline comorbidity | Death, 30 d | 38,381 | 25,582 | 12,799 | 6.04% | 5.36% | 0.68 (0.05 to 1.35) | 1.13 (1.01 to 1.27) |
| No guideline comorbidity | Unplanned readmission, 30 d | 38,381 | 25,582 | 12,799 | 4.92% | 4.36% | 0.56 (0.07 to 0.93) | 1.13 (1.02 to 1.23) |
| No guideline comorbidity | Post-landmark length of stay, h | 38,381 | 25,582 | 12,799 | 112.7 h | 102.3 h | 10.4 (2.9 to 16.0) h | 1.10 (1.03 to 1.16) |
| Guideline comorbidity | Any adverse clinical event, 30 d | 191,782 | 141,172 | 50,610 | 33.26% | 30.96% | 2.30 (1.69 to 3.02) | 1.07 (1.05 to 1.10) |
| Guideline comorbidity | Clinical deterioration, 30 d | 191,782 | 141,172 | 50,610 | 9.68% | 8.13% | 1.54 (1.13 to 1.97) | 1.19 (1.14 to 1.25) |
| Guideline comorbidity | Death, 30 d | 191,782 | 141,172 | 50,610 | 14.03% | 11.99% | 2.04 (1.55 to 2.56) | 1.17 (1.13 to 1.22) |
| Guideline comorbidity | Unplanned readmission, 30 d | 191,782 | 141,172 | 50,610 | 11.29% | 11.17% | 0.12 (-0.22 to 0.45) | 1.01 (0.98 to 1.04) |
| Guideline comorbidity | Post-landmark length of stay, h | 191,782 | 141,172 | 50,610 | 140.9 h | 129.9 h | 11.0 (7.2 to 14.5) h | 1.08 (1.05 to 1.11) |

The sensitivity exposure compared CAP-active administration continued during 48–72 hours with no CAP-active administration during 48–72 hours. CAP = community-acquired pneumonia; CI = confidence interval.

**Supplementary Table S5. Respiratory virus distribution in the primary cohorts**

| **Virus subgroup** | **Inpatient: no guideline comorbidity (N = 47,110)** | **Inpatient: guideline comorbidity (N = 228,494)** | **Outpatient: no guideline comorbidity (N = 50,060)** | **Outpatient: guideline comorbidity (N = 50,656)** |
| --- | --- | --- | --- | --- |
| SARS-CoV-2 | 40,248 (85.4%) | 161,292 (70.6%) | 37,060 (74.0%) | 36,610 (72.3%) |
| Influenza | 3,358 (7.1%) | 28,058 (12.3%) | 7,793 (15.6%) | 6,998 (13.8%) |
| Respiratory syncytial virus | 1,102 (2.3%) | 13,935 (6.1%) | 2,237 (4.5%) | 3,481 (6.9%) |
| Rhinovirus/enterovirus | 1,158 (2.5%) | 12,824 (5.6%) | 1,270 (2.5%) | 1,633 (3.2%) |
| Human metapneumovirus | 530 (1.1%) | 5,313 (2.3%) | 846 (1.7%) | 957 (1.9%) |
| Endemic coronavirus | 313 (0.7%) | 4,231 (1.9%) | 322 (0.6%) | 490 (1.0%) |
| Multiple respiratory viruses | 270 (0.6%) | 2,308 (1.0%) | 375 (0.7%) | 405 (0.8%) |
| Other respiratory virus | 131 (0.3%) | 533 (0.2%) | 157 (0.3%) | 82 (0.2%) |

Values are n (%). Multiple respiratory viruses indicates more than one positive respiratory viral target. Other respiratory virus includes sparse single-virus groups, principally adenovirus.

**Supplementary Table S6. Global treatment-by-virus heterogeneity tests**

| **Analysis** | **Setting** | **Clinical stratum** | **Outcome** | **Eligible virus groups, n** | **Wald χ² (df)** | **P value** |
| --- | --- | --- | --- | --- | --- | --- |
| 72-h sensitivity | Inpatient | No guideline comorbidity | Any adverse clinical event, 30 d | 4 | 6.88 (3) | .0758 |
| 72-h sensitivity | Inpatient | No guideline comorbidity | Post-landmark length of stay | 4 | 4.88 (3) | .1809 |
| 72-h sensitivity | Inpatient | Guideline comorbidity | Any adverse clinical event, 30 d | 6 | 82.66 (5) | <.001 |
| 72-h sensitivity | Inpatient | Guideline comorbidity | Post-landmark length of stay | 6 | 1.58 (5) | .9035 |
| Primary | Inpatient | No guideline comorbidity | Any adverse clinical event, 30 d | 4 | 15.32 (3) | .0016 |
| Primary | Inpatient | No guideline comorbidity | Post-landmark length of stay | 5 | 5.14 (4) | .2735 |
| Primary | Inpatient | Guideline comorbidity | Any adverse clinical event, 30 d | 6 | 83.02 (5) | <.001 |
| Primary | Inpatient | Guideline comorbidity | Death, 30 d | 6 | 74.42 (5) | <.001 |
| Primary | Inpatient | Guideline comorbidity | Post-landmark length of stay | 6 | 3.32 (5) | .6511 |
| 24-h transition sensitivity | Outpatient | No guideline comorbidity | Any adverse clinical event, 30 d | 6 | 28.76 (5) | <.001 |
| 24-h transition sensitivity | Outpatient | No guideline comorbidity | Hospitalization after 24 h | 5 | 17.63 (4) | .0015 |
| 24-h transition sensitivity | Outpatient | Guideline comorbidity | Any adverse clinical event, 30 d | 6 | 30.39 (5) | <.001 |
| 24-h transition sensitivity | Outpatient | Guideline comorbidity | Death, 30 d | 3 | 28.82 (2) | <.001 |
| 24-h transition sensitivity | Outpatient | Guideline comorbidity | Hospitalization after 24 h | 6 | 75.17 (5) | <.001 |

Exploratory Wald tests evaluate whether the overlap-weighted treatment association is constant across eligible virus groups. Covariance was estimated from 500 paired source-cluster bootstrap replications. Eligibility required at least 300 episodes, 50 episodes in each treatment arm, five source clusters, and, for binary outcomes, 10 events in each arm. h = hours.

**Supplementary Table S7. Virus-specific treatment estimates (Part A: primary inpatient adverse-event composite)**

| **Clinical stratum** | **Virus** | **Total, n** | **Continued, n** | **Not continued, n** | **Adjusted risk difference, pp (95% CI)** |
| --- | --- | --- | --- | --- | --- |
| No guideline comorbidity | SARS-CoV-2 | 40,248 | 27,256 | 12,992 | 2.02 (1.26 to 2.83) |
| No guideline comorbidity | Influenza | 3,358 | 3,043 | 315 | -1.02 (-5.34 to 3.04) |
| No guideline comorbidity | Respiratory syncytial virus | 1,102 | 956 | 146 | -6.07 (-13.32 to 0.35) |
| No guideline comorbidity | Rhinovirus/enterovirus | 1,158 | 1,065 | 93 | -8.48 (-15.63 to -1.58) |
| Guideline comorbidity | SARS-CoV-2 | 161,292 | 117,300 | 43,992 | 3.57 (2.92 to 4.32) |
| Guideline comorbidity | Influenza | 28,058 | 24,936 | 3,122 | -0.76 (-2.37 to 0.66) |
| Guideline comorbidity | Respiratory syncytial virus | 13,935 | 11,956 | 1,979 | -2.29 (-4.61 to -0.14) |
| Guideline comorbidity | Rhinovirus/enterovirus | 12,824 | 11,431 | 1,393 | -4.21 (-6.74 to -1.51) |
| Guideline comorbidity | Human metapneumovirus | 5,313 | 4,476 | 837 | 0.71 (-2.55 to 3.64) |
| Guideline comorbidity | Endemic coronavirus | 4,231 | 3,789 | 442 | -0.81 (-4.84 to 3.30) |

Positive values indicate higher 30-day adverse-event risk with CAP-active continuation during hours 24–48. Estimates are shown only for eligible virus–stratum combinations. CI = confidence interval; pp = percentage points.

**Supplementary Table S7. Virus-specific treatment estimates (Part B: inpatient 72-hour adverse-event composite)**

| **Clinical stratum** | **Virus** | **Total, n** | **Continued, n** | **Not continued, n** | **Adjusted risk difference, pp (95% CI)** |
| --- | --- | --- | --- | --- | --- |
| No guideline comorbidity | SARS-CoV-2 | 33,943 | 21,711 | 12,232 | 2.01 (1.13 to 2.78) |
| No guideline comorbidity | Influenza | 2,238 | 1,991 | 247 | -2.11 (-7.19 to 3.09) |
| No guideline comorbidity | Respiratory syncytial virus | 739 | 622 | 117 | 1.03 (-5.52 to 7.38) |
| No guideline comorbidity | Rhinovirus/enterovirus | 705 | 618 | 87 | -6.41 (-14.96 to 1.16) |
| Guideline comorbidity | SARS-CoV-2 | 139,966 | 96,814 | 43,152 | 3.28 (2.54 to 3.89) |
| Guideline comorbidity | Influenza | 21,774 | 19,004 | 2,770 | -1.62 (-3.26 to -0.09) |
| Guideline comorbidity | Respiratory syncytial virus | 11,186 | 9,345 | 1,841 | -2.69 (-4.75 to -0.88) |
| Guideline comorbidity | Rhinovirus/enterovirus | 9,423 | 8,194 | 1,229 | -2.75 (-5.32 to 0.05) |
| Guideline comorbidity | Human metapneumovirus | 4,022 | 3,261 | 761 | 0.21 (-3.95 to 4.19) |
| Guideline comorbidity | Endemic coronavirus | 3,191 | 2,765 | 426 | -1.19 (-5.13 to 2.45) |

The sensitivity exposure compared continuation versus noncontinuation during hours 48–72. Positive values indicate higher 30-day adverse-event risk with continuation. CI = confidence interval; pp = percentage points.

**Supplementary Table S7. Virus-specific treatment estimates (Part C: primary inpatient 30-day mortality with guideline comorbidity)**

| **Virus** | **Total, n** | **Continued, n** | **Not continued, n** | **Adjusted risk difference, pp (95% CI)** |
| --- | --- | --- | --- | --- |
| SARS-CoV-2 | 161,292 | 117,300 | 43,992 | 2.77 (2.21 to 3.33) |
| Influenza | 28,058 | 24,936 | 3,122 | 0.22 (-0.50 to 0.93) |
| Respiratory syncytial virus | 13,935 | 11,956 | 1,979 | -0.43 (-1.58 to 0.73) |
| Rhinovirus/enterovirus | 12,824 | 11,431 | 1,393 | 0.09 (-1.02 to 1.21) |
| Human metapneumovirus | 5,313 | 4,476 | 837 | -0.13 (-1.66 to 1.36) |
| Endemic coronavirus | 4,231 | 3,789 | 442 | 0.07 (-2.58 to 2.60) |

Positive values indicate higher 30-day mortality with CAP-active continuation during hours 24–48. CI = confidence interval; pp = percentage points.

**Supplementary Table S7. Virus-specific treatment estimates (Part D: outpatient 24-hour care-transition adverse-event sensitivity analysis)**

| **Clinical stratum** | **Virus** | **Total, n** | **Prescribed, n** | **Not prescribed, n** | **Adjusted risk difference, pp (95% CI)** |
| --- | --- | --- | --- | --- | --- |
| No guideline comorbidity | SARS-CoV-2 | 37,060 | 14,807 | 22,253 | -2.17 (-3.21 to -1.19) |
| No guideline comorbidity | Influenza | 7,793 | 5,937 | 1,856 | -7.42 (-9.20 to -5.65) |
| No guideline comorbidity | Respiratory syncytial virus | 2,237 | 1,618 | 619 | -4.34 (-7.38 to -1.20) |
| No guideline comorbidity | Rhinovirus/enterovirus | 1,270 | 891 | 379 | -1.27 (-6.65 to 4.16) |
| No guideline comorbidity | Human metapneumovirus | 846 | 452 | 394 | -1.07 (-5.62 to 3.49) |
| No guideline comorbidity | Endemic coronavirus | 322 | 219 | 103 | -12.14 (-22.97 to -2.66) |
| Guideline comorbidity | SARS-CoV-2 | 36,610 | 11,203 | 25,407 | -11.79 (-13.20 to -10.42) |
| Guideline comorbidity | Influenza | 6,998 | 3,792 | 3,206 | -17.25 (-19.47 to -15.23) |
| Guideline comorbidity | Respiratory syncytial virus | 3,481 | 1,811 | 1,670 | -19.00 (-23.00 to -15.52) |
| Guideline comorbidity | Rhinovirus/enterovirus | 1,633 | 900 | 733 | -9.83 (-12.67 to -6.64) |
| Guideline comorbidity | Human metapneumovirus | 957 | 390 | 567 | -7.71 (-11.03 to -4.49) |
| Guideline comorbidity | Endemic coronavirus | 490 | 259 | 231 | -8.35 (-13.94 to -3.12) |

Hospitalizations beginning within 24 hours of emergency-department departure were excluded from the composite. Negative values indicate lower risk with a CAP-active discharge prescription. CI = confidence interval; pp = percentage points.

**Supplementary Table S8. Target-concordant antiviral use in the primary cohorts**

| **Setting** | **Clinical stratum** | **Virus** | **Antibacterial strategy** | **N** | **Target-concordant antiviral, n (%)** |
| --- | --- | --- | --- | --- | --- |
| Inpatient | No guideline comorbidity | SARS-CoV-2 | Continued | 27,256 | 11,986 (44.0%) |
| Inpatient | No guideline comorbidity | SARS-CoV-2 | Not continued | 12,992 | 6,120 (47.1%) |
| Inpatient | No guideline comorbidity | Influenza | Continued | 3,043 | 2,071 (68.1%) |
| Inpatient | No guideline comorbidity | Influenza | Not continued | 315 | 214 (67.9%) |
| Inpatient | Guideline comorbidity | SARS-CoV-2 | Continued | 117,300 | 46,575 (39.7%) |
| Inpatient | Guideline comorbidity | SARS-CoV-2 | Not continued | 43,992 | 19,337 (44.0%) |
| Inpatient | Guideline comorbidity | Influenza | Continued | 24,936 | 15,279 (61.3%) |
| Inpatient | Guideline comorbidity | Influenza | Not continued | 3,122 | 1,666 (53.4%) |
| Outpatient | No guideline comorbidity | SARS-CoV-2 | Prescribed | 14,807 | 850 (5.7%) |
| Outpatient | No guideline comorbidity | SARS-CoV-2 | Not prescribed | 22,253 | 565 (2.5%) |
| Outpatient | No guideline comorbidity | Influenza | Prescribed | 5,937 | 1,723 (29.0%) |
| Outpatient | No guideline comorbidity | Influenza | Not prescribed | 1,856 | 284 (15.3%) |
| Outpatient | Guideline comorbidity | SARS-CoV-2 | Prescribed | 11,203 | 1,092 (9.7%) |
| Outpatient | Guideline comorbidity | SARS-CoV-2 | Not prescribed | 25,407 | 894 (3.5%) |
| Outpatient | Guideline comorbidity | Influenza | Prescribed | 3,792 | 1,261 (33.3%) |
| Outpatient | Guideline comorbidity | Influenza | Not prescribed | 3,206 | 427 (13.3%) |

For inpatients, target-concordant antiviral use was documented before the antibacterial continuation decision. For outpatients, antiviral and antibacterial prescriptions were concurrent at emergency-department discharge.

**Supplementary Table S9. Antiviral-adjusted inpatient sensitivity estimates (Part A: no guideline comorbidity)**

| **Virus** | **Outcome** | **N** | **Continued / not continued weighted estimate** | **Adjusted difference (95% CI)** | **Adjusted ratio (95% CI)** |
| --- | --- | --- | --- | --- | --- |
| SARS-CoV-2 | Any adverse clinical event, 30 d | 40,248 | 18.566% / 16.572% | 1.995 (1.163 to 2.735) | 1.120 (1.068 to 1.170) |
| SARS-CoV-2 | Death, 30 d | 40,248 | 5.998% / 5.369% | 0.629 (0.004 to 1.316) | 1.117 (1.001 to 1.231) |
| SARS-CoV-2 | Post-landmark length of stay | 40,248 | 124.0 h / 107.3 h | 16.7 h (11.3 to 24.6) | 1.156 (1.105 to 1.228) |
| Influenza | Any adverse clinical event, 30 d | 3,358 | 13.357% / 14.389% | -1.031 (-4.960 to 3.116) | 0.928 (0.720 to 1.279) |
| Influenza | Post-landmark length of stay | 3,358 | 66.5 h / 66.6 h | -0.2 h (-11.8 to 9.2) | 0.997 (0.847 to 1.155) |

The virus-specific propensity model additionally included target-concordant antiviral treatment documented before the antibacterial continuation decision. Ratios are risk ratios for binary outcomes and mean ratios for length of stay. CI = confidence interval.

**Supplementary Table S9. Antiviral-adjusted inpatient sensitivity estimates (Part B: guideline comorbidity)**

| **Virus** | **Outcome** | **N** | **Continued / not continued weighted estimate** | **Adjusted difference (95% CI)** | **Adjusted ratio (95% CI)** |
| --- | --- | --- | --- | --- | --- |
| SARS-CoV-2 | Any adverse clinical event, 30 d | 161,292 | 35.259% / 31.930% | 3.329 (2.563 to 4.035) | 1.104 (1.080 to 1.128) |
| SARS-CoV-2 | Death, 30 d | 161,292 | 15.540% / 12.943% | 2.597 (2.041 to 3.173) | 1.201 (1.153 to 1.253) |
| SARS-CoV-2 | Post-landmark length of stay | 161,292 | 159.1 h / 147.1 h | 12.0 h (6.6 to 16.3) | 1.082 (1.043 to 1.113) |
| Influenza | Any adverse clinical event, 30 d | 28,058 | 23.178% / 23.847% | -0.674 (-2.208 to 1.146) | 0.972 (0.912 to 1.051) |
| Influenza | Death, 30 d | 28,058 | 4.758% / 4.455% | 0.303 (-0.520 to 0.963) | 1.068 (0.899 to 1.245) |
| Influenza | Post-landmark length of stay | 28,058 | 95.0 h / 85.9 h | 9.1 h (3.4 to 16.0) | 1.106 (1.038 to 1.193) |

The virus-specific propensity model additionally included target-concordant antiviral treatment documented before the antibacterial continuation decision. Ratios are risk ratios for binary outcomes and mean ratios for length of stay. CI = confidence interval.

**Supplementary Table S10. Antibacterial regimen distributions among treated patients (Part A: inpatients)**

| **Regimen category** | **No comorbidity: SARS-CoV-2 (N = 27,256)** | **No comorbidity: influenza (N = 3,043)** | **Comorbidity: SARS-CoV-2 (N = 117,300)** | **Comorbidity: influenza (N = 24,936)** |
| --- | --- | --- | --- | --- |
| β-lactam plus macrolide | 12,370 (45.4%) | 1,200 (39.4%) | 41,536 (35.4%) | 8,211 (32.9%) |
| β-lactam plus tetracycline | 2,775 (10.2%) | 411 (13.5%) | 13,783 (11.8%) | 3,504 (14.1%) |
| β-lactam alone | 3,103 (11.4%) | 348 (11.4%) | 13,718 (11.7%) | 2,472 (9.9%) |
| Macrolide alone | 2,941 (10.8%) | 141 (4.6%) | 10,065 (8.6%) | 1,529 (6.1%) |
| Tetracycline-based | 1,560 (5.7%) | 69 (2.3%) | 6,787 (5.8%) | 1,056 (4.2%) |
| Respiratory fluoroquinolone | 1,038 (3.8%) | 156 (5.1%) | 3,877 (3.3%) | 1,137 (4.6%) |
| Broad anti-MRSA or antipseudomonal | 3,365 (12.3%) | 690 (22.7%) | 26,964 (23.0%) | 6,853 (27.5%) |
| Other or multiclass CAP-active | 104 (0.4%) | 26 (0.9%) | 570 (0.5%) | 174 (0.7%) |

Values are n (%). Regimen categories were assigned from CAP-active medications during the exposure window. The table is descriptive; regimen choice is part of treatment and is confounded by indication. MRSA = methicillin-resistant Staphylococcus aureus.

**Supplementary Table S10. Antibacterial regimen distributions among treated patients (Part B: outpatients)**

| **Regimen category** | **No comorbidity: SARS-CoV-2 (N = 14,807)** | **No comorbidity: influenza (N = 5,937)** | **Comorbidity: SARS-CoV-2 (N = 11,203)** | **Comorbidity: influenza (N = 3,792)** |
| --- | --- | --- | --- | --- |
| β-lactam plus macrolide | 1,933 (13.1%) | 1,162 (19.6%) | 1,622 (14.5%) | 756 (19.9%) |
| β-lactam plus tetracycline | 714 (4.8%) | 509 (8.6%) | 878 (7.8%) | 444 (11.7%) |
| β-lactam alone | 1,315 (8.9%) | 838 (14.1%) | 1,140 (10.2%) | 465 (12.3%) |
| Macrolide alone | 6,643 (44.9%) | 1,451 (24.4%) | 4,031 (36.0%) | 819 (21.6%) |
| Tetracycline-based | 3,437 (23.2%) | 1,532 (25.8%) | 2,723 (24.3%) | 910 (24.0%) |
| Respiratory fluoroquinolone | 618 (4.2%) | 387 (6.5%) | 666 (5.9%) | 352 (9.3%) |
| Broad anti-MRSA or antipseudomonal | 1 (<0.1%) | 0 | 3 (<0.1%) | 1 (<0.1%) |
| Other or multiclass CAP-active | 146 (1.0%) | 58 (1.0%) | 140 (1.2%) | 45 (1.2%) |

Values are n (%). Regimen categories were assigned from CAP-active prescriptions at emergency-department discharge. The table is descriptive; regimen choice is part of treatment and is confounded by indication. MRSA = methicillin-resistant Staphylococcus aureus.

**Supplementary Table S11. Exploratory inpatient antibacterial-class continuation analyses**

| **Analysis** | **Outcome** | **Clinical stratum** | **Class/regimen contrast** | **Continued / not continued, n** | **Adjusted risk difference, pp (95% CI)** |
| --- | --- | --- | --- | --- | --- |
| Primary 24–48 h | Any adverse clinical event, 30 d | No guideline comorbidity | Any atypical coverage | 26,855 / 13,703 | 0.12 (-0.73 to 0.88) |
| Primary 24–48 h | Any adverse clinical event, 30 d | No guideline comorbidity | Macrolide-containing | 19,402 / 13,703 | 0.13 (-0.75 to 0.97) |
| Primary 24–48 h | Any adverse clinical event, 30 d | No guideline comorbidity | Doxycycline/tetracycline-containing | 6,048 / 13,703 | -0.25 (-1.87 to 1.12) |
| Primary 24–48 h | Any adverse clinical event, 30 d | No guideline comorbidity | Respiratory fluoroquinolone | 1,643 / 13,703 | 0.38 (-2.09 to 2.53) |
| Primary 24–48 h | Any adverse clinical event, 30 d | No guideline comorbidity | Beta-lactam without atypical coverage | 6,208 / 13,703 | 3.58 (2.12 to 4.84) |
| Primary 24–48 h | Any adverse clinical event, 30 d | No guideline comorbidity | Broad anti-MRSA/antipseudomonal | 4,734 / 13,703 | 6.89 (5.28 to 8.63) |
| Primary 24–48 h | Any adverse clinical event, 30 d | Guideline comorbidity | Any atypical coverage | 129,932 / 52,219 | -0.03 (-0.65 to 0.61) |
| Primary 24–48 h | Any adverse clinical event, 30 d | Guideline comorbidity | Macrolide-containing | 84,538 / 52,219 | -0.80 (-1.54 to -0.10) |
| Primary 24–48 h | Any adverse clinical event, 30 d | Guideline comorbidity | Doxycycline/tetracycline-containing | 38,137 / 52,219 | 1.02 (0.16 to 1.99) |
| Primary 24–48 h | Any adverse clinical event, 30 d | Guideline comorbidity | Respiratory fluoroquinolone | 8,636 / 52,219 | 1.43 (0.15 to 2.77) |
| Primary 24–48 h | Any adverse clinical event, 30 d | Guideline comorbidity | Beta-lactam without atypical coverage | 44,370 / 52,219 | 4.73 (3.94 to 5.61) |
| Primary 24–48 h | Any adverse clinical event, 30 d | Guideline comorbidity | Broad anti-MRSA/antipseudomonal | 42,386 / 52,219 | 7.79 (6.84 to 8.65) |
| Primary 24–48 h | Death, 30 d | No guideline comorbidity | Any atypical coverage | 26,855 / 13,703 | -0.53 (-1.13 to 0.06) |
| Primary 24–48 h | Death, 30 d | No guideline comorbidity | Macrolide-containing | 19,402 / 13,703 | -0.45 (-1.00 to 0.12) |
| Primary 24–48 h | Death, 30 d | No guideline comorbidity | Doxycycline/tetracycline-containing | 6,048 / 13,703 | -0.90 (-2.05 to -0.03) |
| Primary 24–48 h | Death, 30 d | No guideline comorbidity | Respiratory fluoroquinolone | 1,643 / 13,703 | -0.62 (-1.90 to 0.50) |
| Primary 24–48 h | Death, 30 d | No guideline comorbidity | Beta-lactam without atypical coverage | 6,208 / 13,703 | 1.28 (0.34 to 2.19) |
| Primary 24–48 h | Death, 30 d | No guideline comorbidity | Broad anti-MRSA/antipseudomonal | 4,734 / 13,703 | 3.08 (1.80 to 4.33) |
| Primary 24–48 h | Death, 30 d | Guideline comorbidity | Any atypical coverage | 129,932 / 52,219 | -0.03 (-0.54 to 0.52) |
| Primary 24–48 h | Death, 30 d | Guideline comorbidity | Macrolide-containing | 84,538 / 52,219 | -0.35 (-0.98 to 0.14) |
| Primary 24–48 h | Death, 30 d | Guideline comorbidity | Doxycycline/tetracycline-containing | 38,137 / 52,219 | 0.50 (-0.20 to 1.23) |
| Primary 24–48 h | Death, 30 d | Guideline comorbidity | Respiratory fluoroquinolone | 8,636 / 52,219 | 0.22 (-0.91 to 1.24) |
| Primary 24–48 h | Death, 30 d | Guideline comorbidity | Beta-lactam without atypical coverage | 44,370 / 52,219 | 3.95 (3.31 to 4.59) |
| Primary 24–48 h | Death, 30 d | Guideline comorbidity | Broad anti-MRSA/antipseudomonal | 42,386 / 52,219 | 6.05 (5.23 to 6.81) |
| 72-h sensitivity | Any adverse clinical event, 30 d | No guideline comorbidity | Any atypical coverage | 19,046 / 12,799 | 0.78 (-0.42 to 1.68) |
| 72-h sensitivity | Any adverse clinical event, 30 d | No guideline comorbidity | Macrolide-containing | 13,169 / 12,799 | 0.80 (-0.26 to 1.70) |
| 72-h sensitivity | Any adverse clinical event, 30 d | No guideline comorbidity | Broad anti-MRSA/antipseudomonal | 3,589 / 12,799 | 9.23 (7.77 to 10.69) |
| 72-h sensitivity | Any adverse clinical event, 30 d | Guideline comorbidity | Any atypical coverage | 95,019 / 50,610 | 0.21 (-0.49 to 0.93) |
| 72-h sensitivity | Any adverse clinical event, 30 d | Guideline comorbidity | Macrolide-containing | 57,422 / 50,610 | -0.92 (-1.64 to -0.17) |
| 72-h sensitivity | Any adverse clinical event, 30 d | Guideline comorbidity | Broad anti-MRSA/antipseudomonal | 33,815 / 50,610 | 10.59 (9.55 to 11.53) |
| 72-h sensitivity | Death, 30 d | No guideline comorbidity | Any atypical coverage | 19,046 / 12,799 | 0.07 (-0.68 to 0.82) |
| 72-h sensitivity | Death, 30 d | No guideline comorbidity | Macrolide-containing | 13,169 / 12,799 | 0.20 (-0.51 to 0.89) |
| 72-h sensitivity | Death, 30 d | No guideline comorbidity | Broad anti-MRSA/antipseudomonal | 3,589 / 12,799 | 5.73 (4.38 to 7.16) |
| 72-h sensitivity | Death, 30 d | Guideline comorbidity | Any atypical coverage | 95,019 / 50,610 | 0.30 (-0.23 to 0.82) |
| 72-h sensitivity | Death, 30 d | Guideline comorbidity | Macrolide-containing | 57,422 / 50,610 | -0.14 (-0.76 to 0.42) |
| 72-h sensitivity | Death, 30 d | Guideline comorbidity | Broad anti-MRSA/antipseudomonal | 33,815 / 50,610 | 8.55 (7.67 to 9.30) |

Each class/regimen indicator was evaluated in a separate exploratory overlap-weighted comparison against the same noncontinuation group within the relevant stratum and landmark. Class indicators overlap and therefore should not be interpreted as mutually exclusive head-to-head treatment groups. The 72-hour replication was limited to the three prespecified contrasts shown. CI = confidence interval; MRSA = methicillin-resistant Staphylococcus aureus; pp = percentage points.

**Supplementary Table S12. Outpatient sensitivity restricted to no captured post-result ED antibacterial administration**

| **Clinical stratum** | **Outcome** | **N (prescription / no prescription)** | **Adjusted risk difference, pp (95% CI)** | **Risk ratio (95% CI)** |
| --- | --- | --- | --- | --- |
| No guideline comorbidity | Any adverse clinical event, 30 d | 50,020 (24,227 / 25,793) | -7.17 (-8.81 to -5.66) | 0.75 (0.70 to 0.79) |
| No guideline comorbidity | Hospitalization, 14 d | 50,020 (24,227 / 25,793) | -8.43 (-10.02 to -6.87) | 0.51 (0.46 to 0.58) |
| No guideline comorbidity | Death, 30 d | 50,020 (24,227 / 25,793) | -0.67 (-0.88 to -0.48) | 0.55 (0.45 to 0.66) |
| No guideline comorbidity | ED revisit, 30 d | 50,020 (24,227 / 25,793) | -1.55 (-2.53 to -0.76) | 0.93 (0.89 to 0.97) |
| No guideline comorbidity | Adverse-event composite excluding hospitalization within 24 h | 50,020 (24,227 / 25,793) | -2.24 (-3.20 to -1.44) | 0.91 (0.86 to 0.94) |
| No guideline comorbidity | Hospitalization after 24 h through day 14 | 50,020 (24,227 / 25,793) | -1.53 (-2.14 to -0.90) | 0.84 (0.78 to 0.91) |
| Guideline comorbidity | Any adverse clinical event, 30 d | 50,633 (18,560 / 32,073) | -21.31 (-23.35 to -19.32) | 0.57 (0.55 to 0.60) |
| Guideline comorbidity | Hospitalization, 14 d | 50,633 (18,560 / 32,073) | -23.54 (-25.97 to -21.29) | 0.37 (0.35 to 0.40) |
| Guideline comorbidity | Death, 30 d | 50,633 (18,560 / 32,073) | -2.01 (-2.41 to -1.60) | 0.57 (0.50 to 0.64) |
| Guideline comorbidity | ED revisit, 30 d | 50,633 (18,560 / 32,073) | -9.80 (-11.03 to -8.58) | 0.74 (0.71 to 0.77) |
| Guideline comorbidity | Adverse-event composite excluding hospitalization within 24 h | 50,633 (18,560 / 32,073) | -11.61 (-12.70 to -10.58) | 0.71 (0.69 to 0.73) |
| Guideline comorbidity | Hospitalization after 24 h through day 14 | 50,633 (18,560 / 32,073) | -6.02 (-7.10 to -5.02) | 0.67 (0.62 to 0.72) |

Episodes with any captured post-result CAP-active antibacterial administration in the emergency department were excluded. For the early-transition-restricted rows, hospitalization beginning within 24 hours of emergency-department departure was excluded from both the hospitalization outcome and the adverse-event composite. Estimates use overlap weighting and 500 source-cluster bootstrap replications. CI = confidence interval; ED = emergency department; pp = percentage points.

**Supplementary Figure S1. Covariate balance: 30 largest baseline imbalances**

The 30 largest pre-weighting absolute standardized mean differences are shown using the same symbols and threshold as Figure 2.

Alt text: The extended Love plot includes 30 covariate indicators. Before weighting, several viral, calendar-year, and source-group terms exceed 0.10; after overlap weighting all displayed terms are essentially zero.

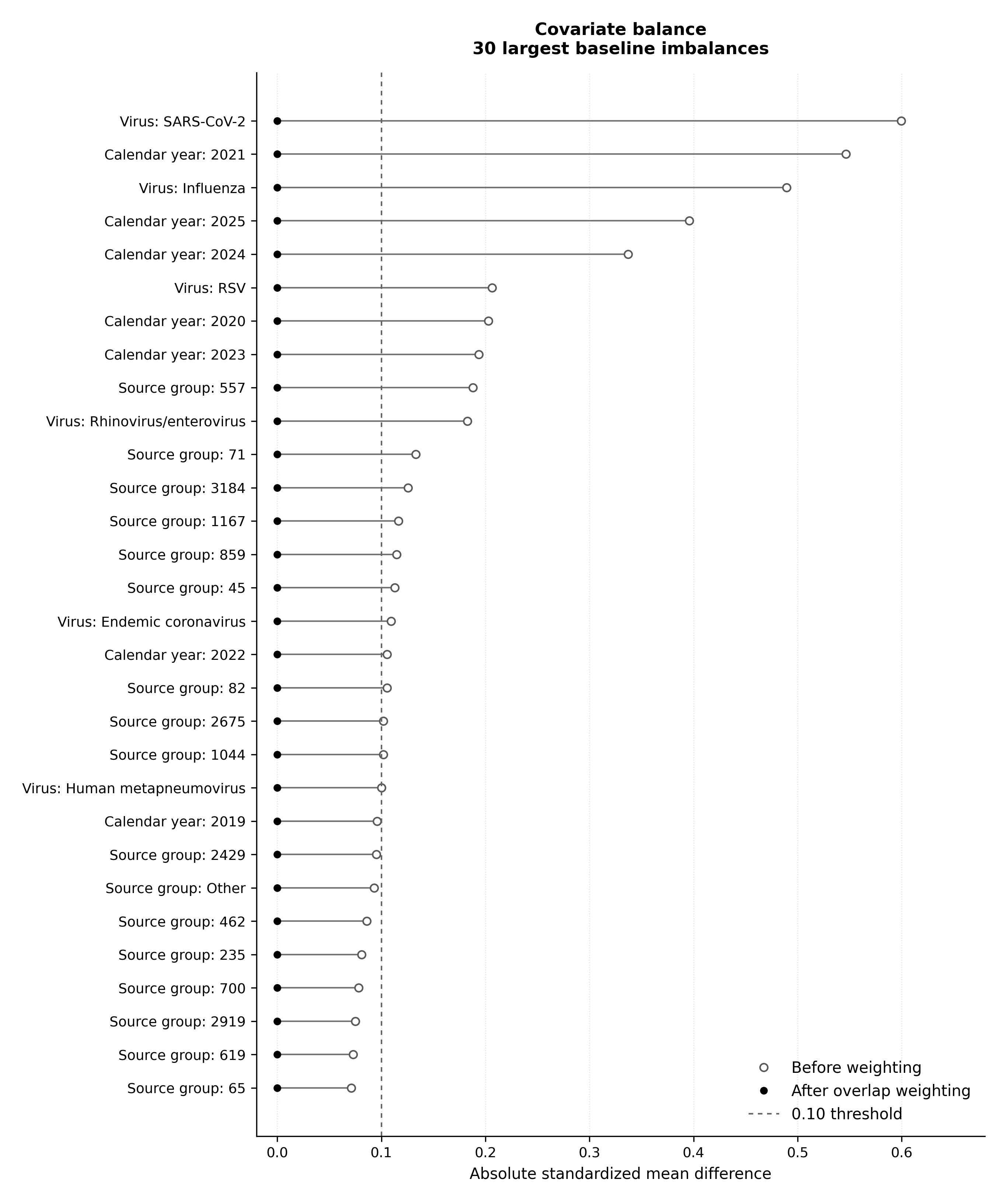

**Supplementary Figure S2. Respiratory virus distribution in the primary cohorts**

Horizontal stacked bars show the percentage distribution of respiratory virus subgroups in the four primary setting and comorbidity strata. SARS-CoV-2 predominates in every stratum.

Alt text: Stacked bar chart of virus distribution in four primary cohorts.

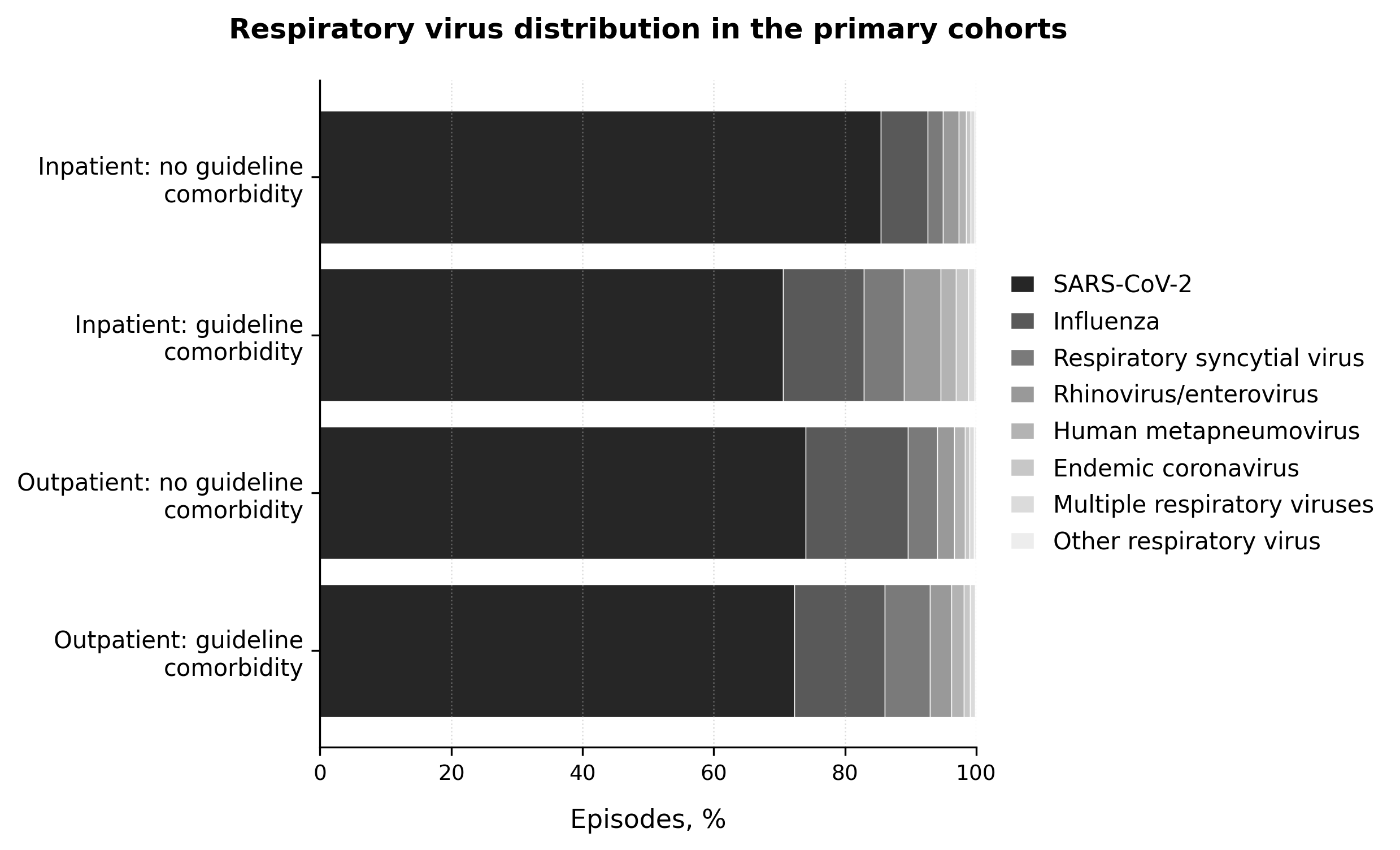

**Supplementary Figure S3. Virus-specific outpatient adverse-event estimates**

Adjusted risk differences compare CAP-active prescription versus no prescription in the 24-hour care-transition sensitivity analysis. Points are overlap-weighted estimates and horizontal lines are 95% source-cluster bootstrap confidence intervals. Negative values indicate lower risk with prescription. Only eligible virus–stratum combinations are shown.

Alt text: Forest plots of outpatient adjusted risk differences by virus and comorbidity stratum.

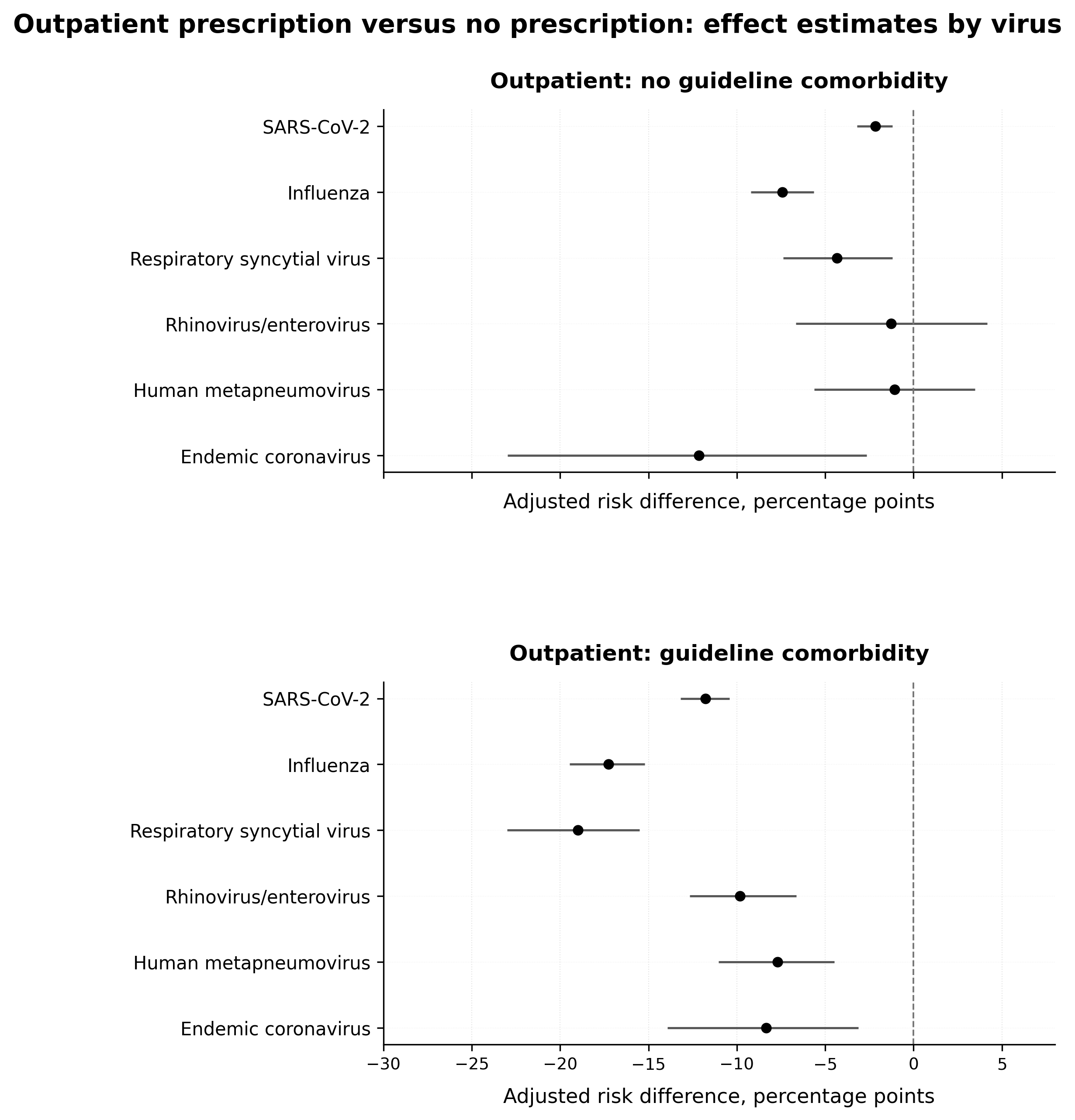

**Supplementary Figure S4. Target-concordant antiviral use in the primary cohorts**

Bars show target-concordant antiviral use for SARS-CoV-2 and influenza, separately by setting, guideline-comorbidity stratum, and antibacterial strategy. Inpatient antivirals were documented before the antibacterial continuation decision; outpatient antiviral and antibacterial prescriptions were concurrent at discharge.

Alt text: Horizontal bar charts of antiviral use for SARS-CoV-2 and influenza.

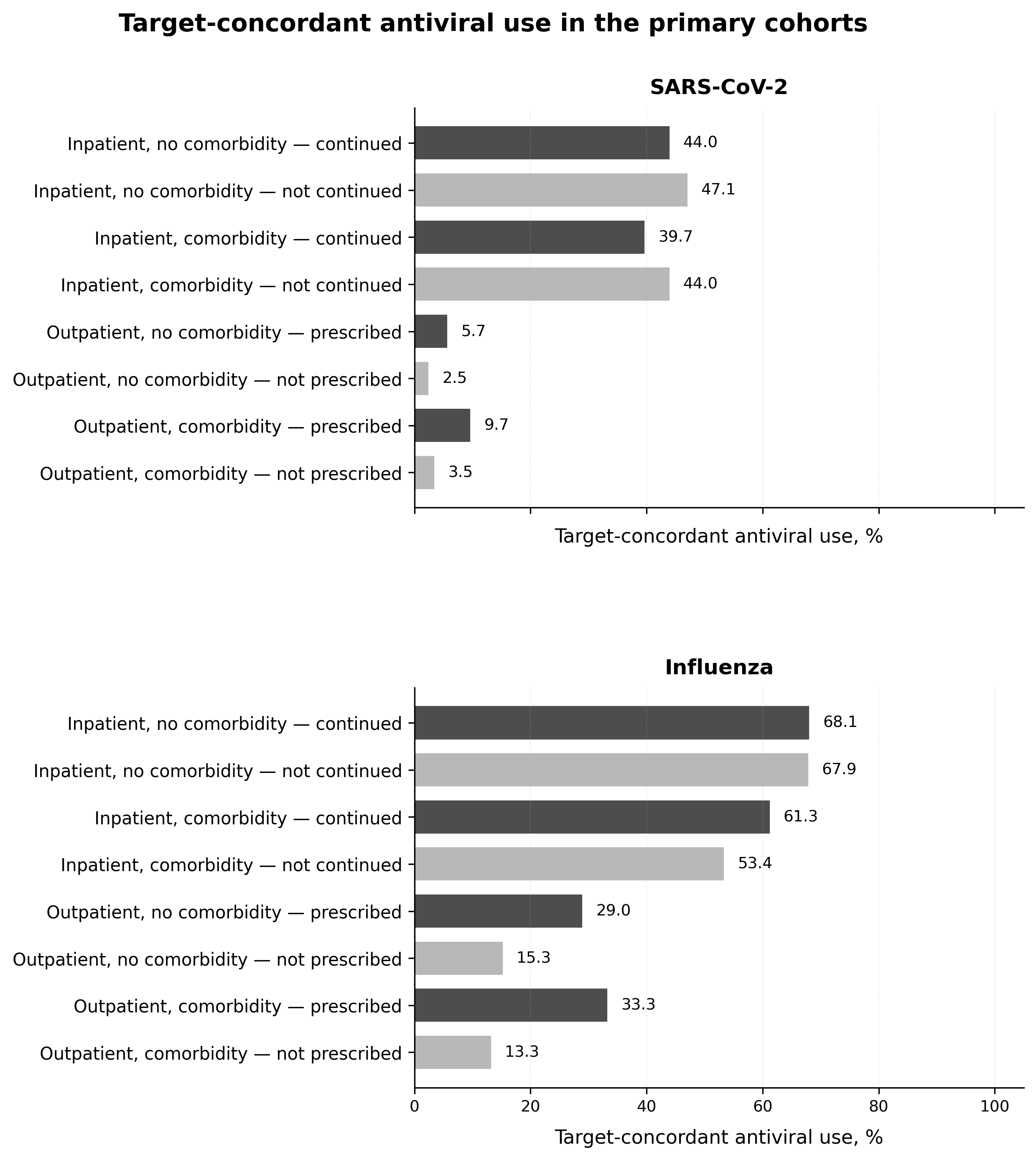

**Supplementary Figure S5A. Primary 24–48-hour inpatient antibacterial-class continuation and any adverse clinical event at 30 days**

Exploratory overlap-weighted adjusted risk differences compare continuation versus noncontinuation of each overlapping antibacterial-class indicator. Points are adjusted risk differences and horizontal lines are 95% source-cluster bootstrap confidence intervals. Positive values indicate higher risk with continuation.

Alt text: Two-panel forest plot for patients without and with guideline comorbidity. Broad anti-MRSA/antipseudomonal therapy and beta-lactam therapy without atypical coverage have the largest positive adverse-event risk differences; atypical and macrolide-containing estimates are near the null.

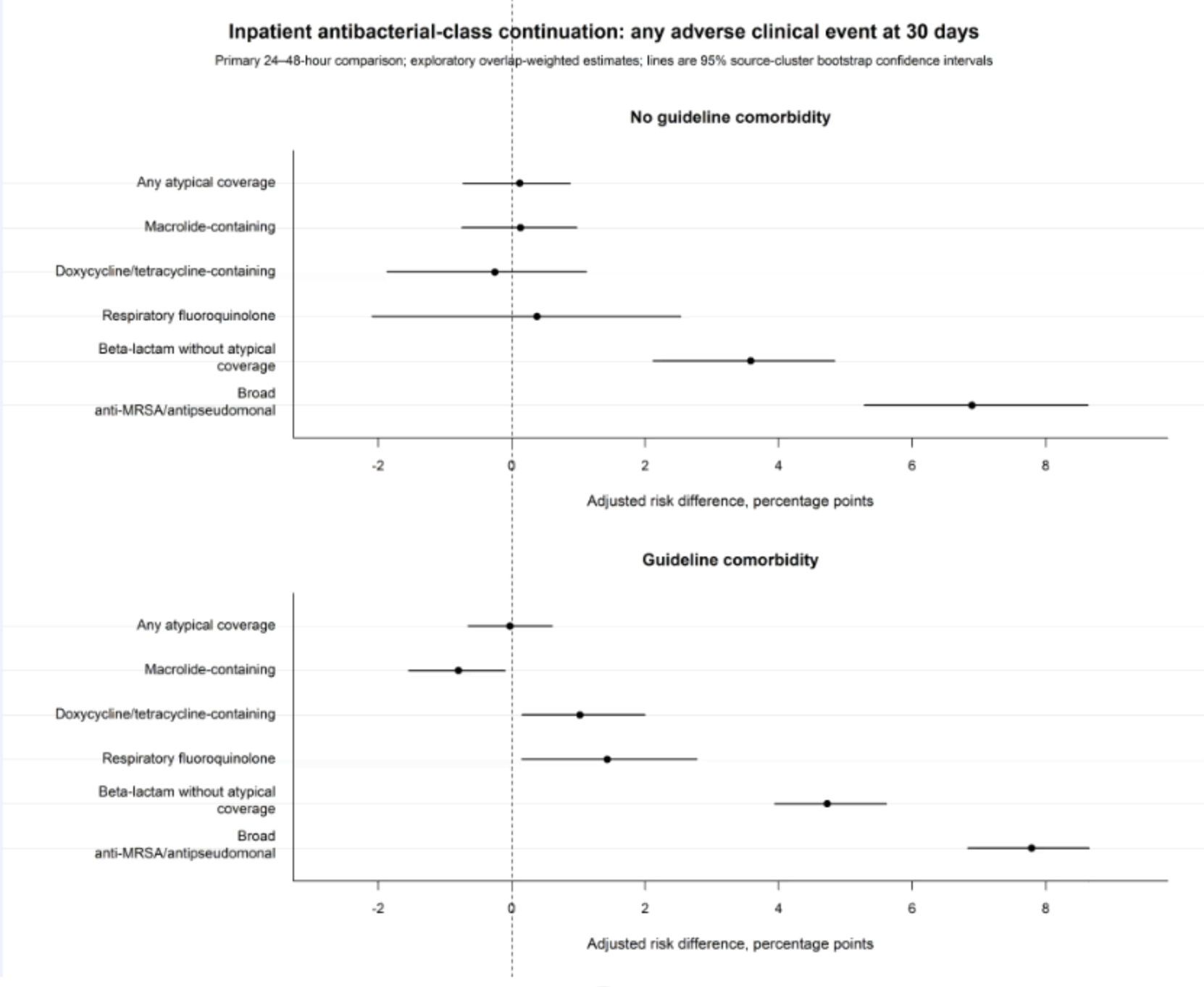

**Supplementary Figure S5B. Primary 24–48-hour inpatient antibacterial-class continuation and death at 30 days**

Exploratory overlap-weighted adjusted risk differences compare continuation versus noncontinuation of each overlapping antibacterial-class indicator. Positive values indicate higher 30-day mortality with continuation.

Alt text: Two-panel forest plot shows positive mortality risk differences for broad anti-MRSA/antipseudomonal and beta-lactam therapy without atypical coverage; estimates for atypical and macrolide-containing therapy are near the null.

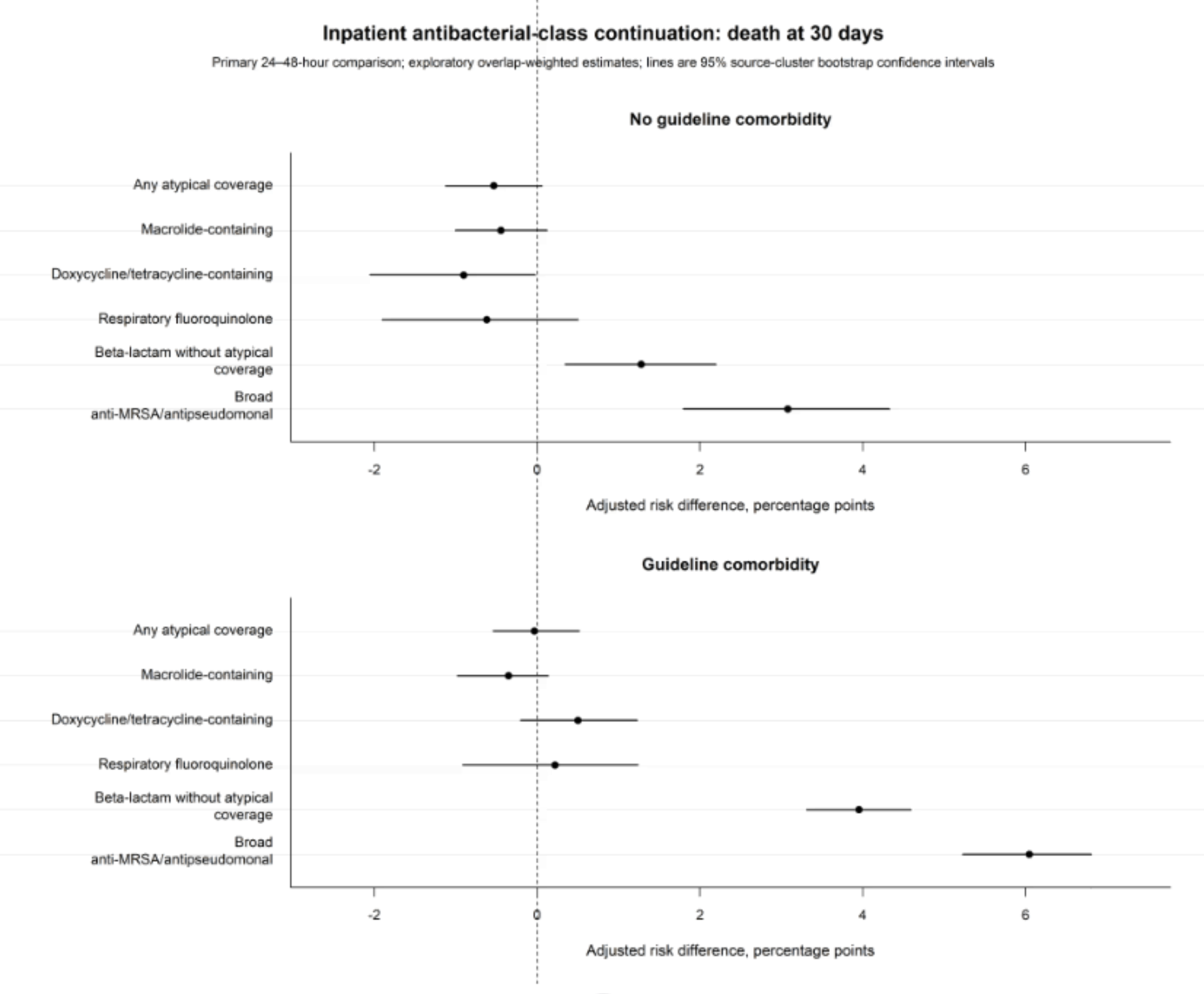

**Supplementary Figure S5C. Inpatient 72-hour antibacterial-class sensitivity analysis and any adverse clinical event at 30 days**

Continuation during hours 48–72 was compared with no continuation for three prespecified class/regimen contrasts. Points are adjusted risk differences and horizontal lines are 95% source-cluster bootstrap confidence intervals.

Alt text: Two-panel forest plot shows large positive adverse-event risk differences for broad anti-MRSA/antipseudomonal therapy and estimates near the null for atypical or macrolide-containing therapy.

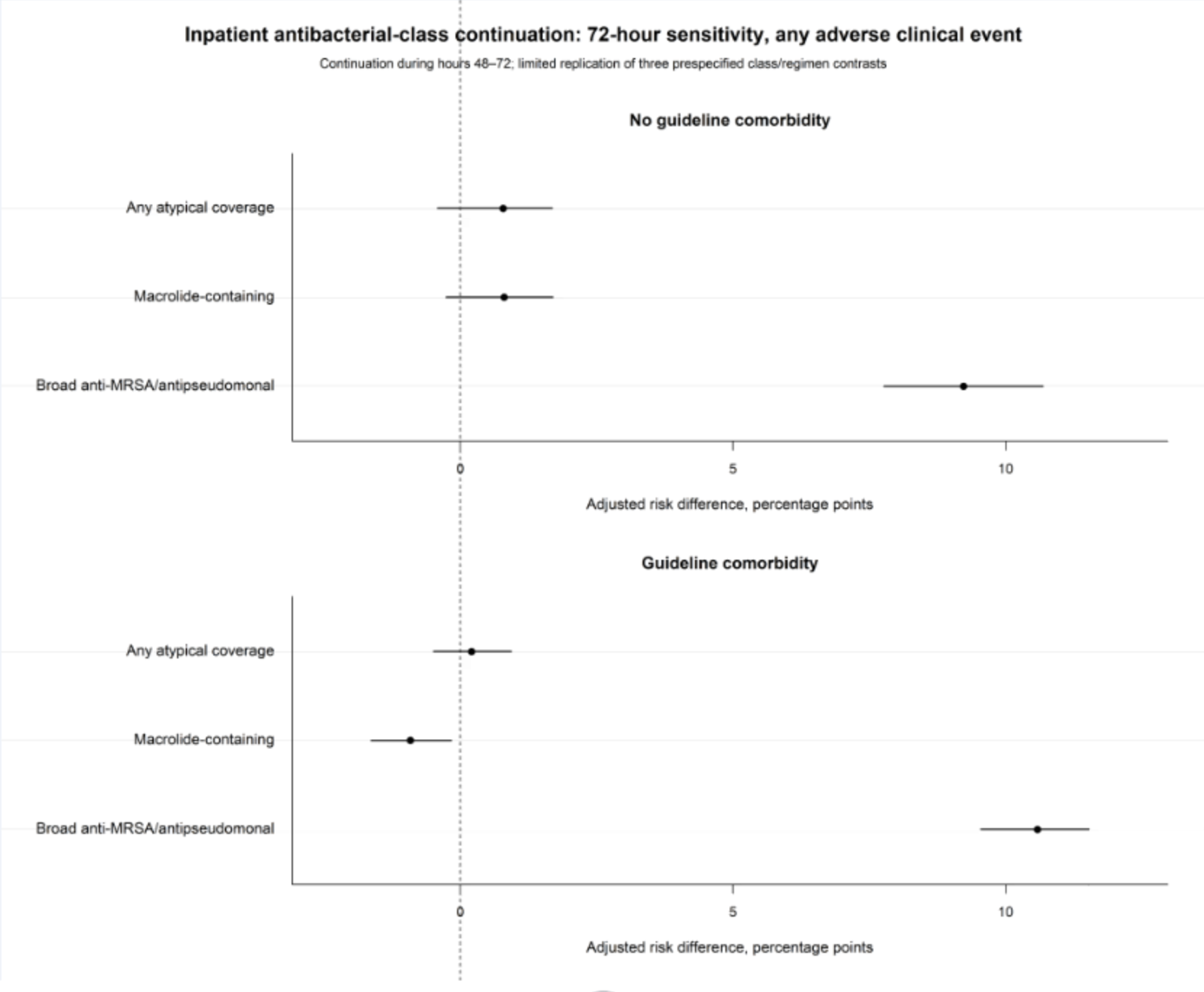

**Supplementary Figure S5D. Inpatient 72-hour antibacterial-class sensitivity analysis and death at 30 days**

Continuation during hours 48–72 was compared with no continuation for three prespecified class/regimen contrasts. Positive values indicate higher 30-day mortality with continuation.

Alt text: Two-panel forest plot shows large positive mortality risk differences for broad anti-MRSA/antipseudomonal therapy and estimates near the null for atypical or macrolide-containing therapy.

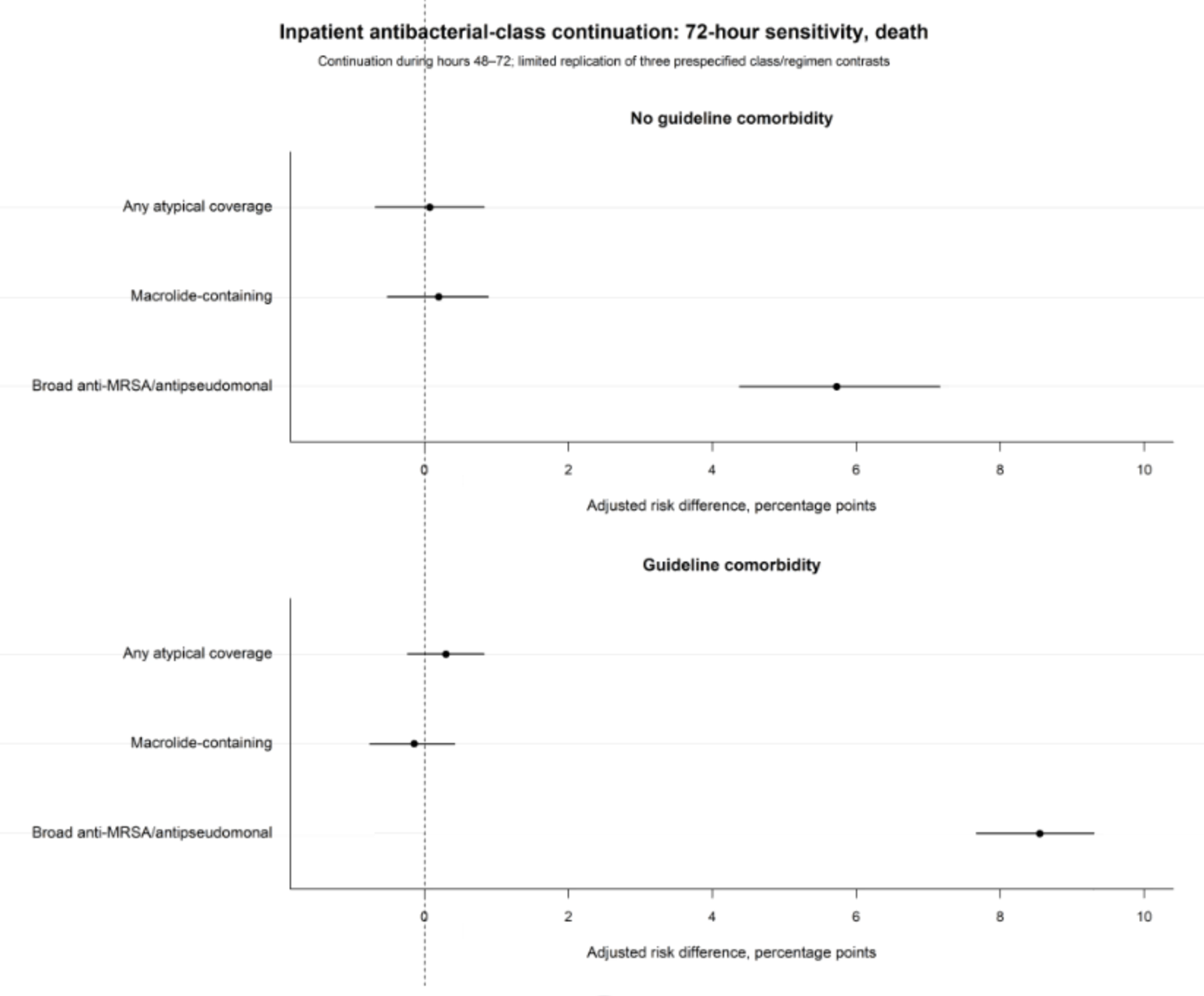

**Supplementary Figure S6. Outpatient prescription association by time to hospitalization**

Adjusted risk differences are shown for hospitalization beginning 0–6 hours, more than 6–24 hours, more than 24–72 hours, and more than 72 hours through day 14 after emergency-department departure. Negative values indicate lower observed risk among patients receiving a discharge prescription.

Alt text: Two-panel forest plot shows the largest negative associations for hospitalization within 6 hours of departure and progressive attenuation toward the null for later hospitalization windows.

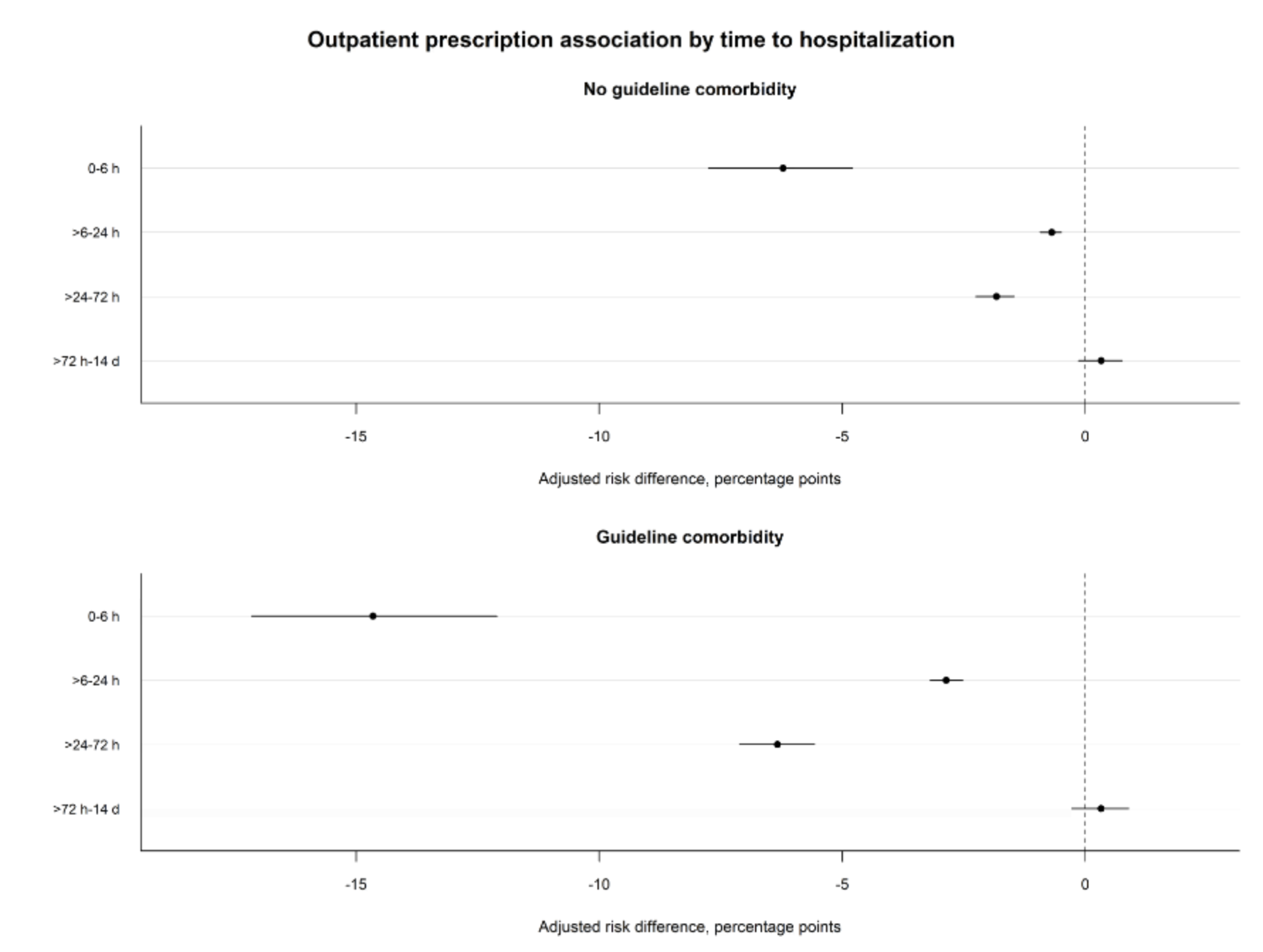

**Supplementary Figure S7. Outpatient sensitivity restricted to no captured post-result ED antibacterial administration**

Prescription versus no prescription was re-estimated after excluding episodes with captured post-result emergency-department antibacterial administration. Early-transition-restricted outcomes also excluded hospitalization beginning within 24 hours.

Alt text: Two-panel forest plot shows lower observed adverse-event and hospitalization risks among prescribed patients after excluding captured post-result emergency-department administration; the estimates attenuate when early hospitalizations are excluded.

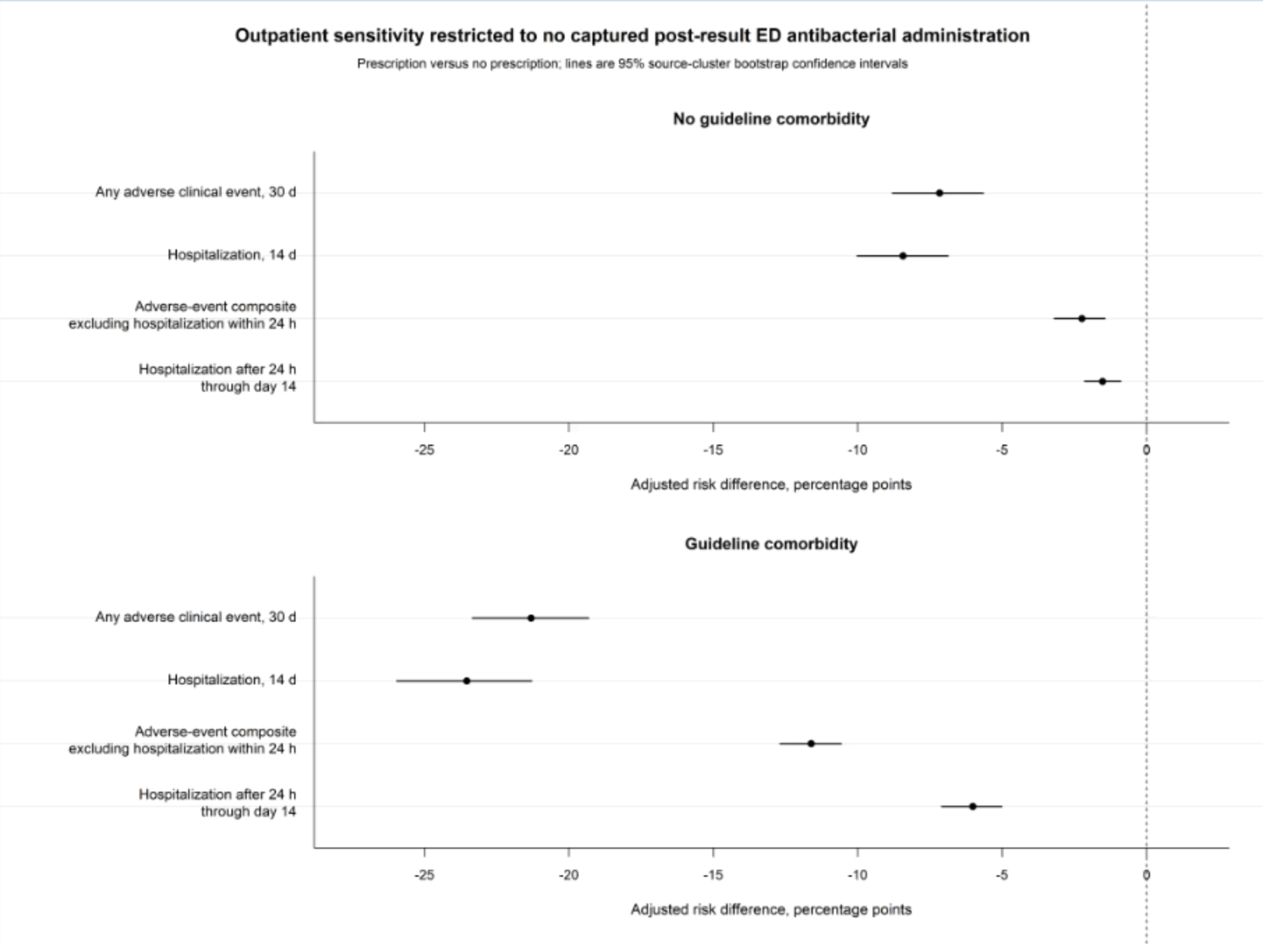
